# Diagnostic utility of in vivo expressed mycobacterial RNA transcripts in pleural fluid for the differential diagnosis of tuberculous pleuritis

**DOI:** 10.1101/2024.01.09.24300747

**Authors:** Prabhdeep Kaur, Sumedha Sharma, Sudhanshu Abhishek Sinha, Ashutosh N. Aggarwal, Khushpreet Kaur, Rakesh Yadav, Sunil Sethi, Amanjit Bal, Indu Verma

## Abstract

**Background:** Tuberculous pleuritis (TBP), the extra-pulmonary manifestation of tuberculosis, is the second most common after TB lymphadenitis. Histopathology using biopsy samples is the most sensitive diagnostic procedure for TBP, however the biopsy acquisition is invasive. Therefore, better screening markers for diagnosis using pleural fluid are required. The pathogen biomarkers expressed at the site of infection may play a potential role in designing a newer diagnostic assay. Thus, the current study was planned to look for mycobacterial RNA biomarkers in TBP and to assess their diagnostic utility in pleural fluid.

**Methods:** TBP suspects (n=261) were recruited in the current study. Out of these 45 suspects were excluded and the remaining (n=216) were divided into TBP (n=54) and non-TBP (n=162) groups based on composite reference standard. A whole genome microarray was carried using *M.tb* RNA from pleural biopsies of TB patients. The data was validated using qRT-PCR and the diagnostic utility of top two highly expressed genes was assessed in pleural fluid of using a real time RT-PCR assay.

**Results:** Overall, 1856 genes were differentially expressed in microarray of which 1365 were upregulated and 491 were downregulated. After validation of microarray gene expression, two genes namely *Rv1586* and *Rv2543* were selected for assessment of their diagnostic utility in TBP. The combined analysis for the presence of either of genes in the pleural fluid led to identification of pleural TB patients with 79.6% sensitivity and 93.28% specificity.

**Conclusion:** The transcripts of genes *Rv1586* and *Rv2543* holds potential for the development of a RNA based molecular diagnostic assay in pleural fluid of TBP patients.

## INTRODUCTION

Tuberculosis typically affects lungs, but can also affect other sites. [1]. Extra-pulmonary TB (EPTB) accounts for approximately 15-20% cases in HIV negative people, and 50% in HIV positive individuals [2]. Tuberculous pleuritis (TB pleuritis/TBP) is the most prevalent form of extra-pulmonary tuberculosis globally after lymph node TB [2,3]. The diagnosis of TB pleuritis is carried out using pleural fluid and biopsy acquired through thoracentesis and thoracoscopy respectively. Considering the paucibacillary nature of TBP, the conventional diagnostic tests including pleural fluid microscopy and culture offer low sensitivity [4–7]. Pleural fluid cytological findings are mostly diagnostic for malignant effusions [2,8,9]. Histopathological examination of pleural biopsy exhibits highest diagnostic potential; however, biopsy acquisition is an invasive procedure [10,11] and associated with post procedure complications [12–14]. The adenosine deaminase (ADA), interferon gamma (IFN-γ) and T cell based IFN-γ release assays (IGRAs) [15–19] are rapid and cost effective, but are nonspecific and the utilization of in-house detection procedures [2,15,18] and lack of a widely accepted cutoff are major constrains limiting their utilization [20]. The utility of Xpert MTB/RIF, the only molecular assay available for TBP is limited [2,21]. The improved version Ultra has better sensitivity and has been endorsed by WHO for TBP diagnosis [22–24]. However, these NAAT’s are based on DNA and cannot differentiate live/dead bacilli which could be reason for false positive Xpert MTB/RIF results in previously treated TB patients [25]. To address these limitations, in the current study, an RNA-based diagnostic test is proposed for TBP. For an RNA based assay the transcripts expressed *in-vivo* at the site of infection by the pathogen could be important targets. Mycobacterial gene expression is known to be altered under different growth conditions as well as in vivo at different sites [26–32]. The study is based on the hypothesis that in vivo highly expressed mycobacterial genes particularly during active disease could be important signatures for the development of a sensitive and specific molecular diagnostic assay. Thus, the present study was carried out to analyse the mycobacterial gene expression profile in biopsy samples of TB pleuritis and identify mycobacterial transcripts that can be utilized as potential candidates for RNA based molecular assay for the diagnosis of pleural TB using pleural fluid samples. Though, RNA based diagnostic assays are difficult to perform due to technical complexities but a widespread use of these assays in the COVID era has provided new hopes for use of such assays for TB diagnosis.

## MATERIALS AND METHODS

### Study subjects

The study subjects for the present study were recruited from Department of Pulmonary Medicine, PGIMER following the approval of study by Institutional Ethics Committee (IEC) (Ref no. INT/IEC/2016/2264). Patients with exudative pleural effusions, history of fever and/or respiratory symptoms for ˃2 weeks and referred for thoracocentesis for diagnostic/therapeutic purpose were recruited as tuberculosis pleuritis (TBP) suspects (n=261) with prior written informed consent. Patients already on anti-tubercular therapy (ATT) at time of recruitment or having positive HIV/HCV status or with any other co-morbidity were excluded from the study.

#### Sample collection

Pleural biopsy and pleural fluid samples were collected from study subjects using thoracoscopy and thoracentesis, respectively by trained physicians. All the samples were also subjected to routine diagnostic tests and the reports were prospectively collected. Clinical and demographic details were collected at the time of enrollment.

#### Diagnostic algorithm for categorization of TBP suspects

A composite reference standard (CRS) comprising of clinical findings (history of fever or respiratory symptoms along with radiological evidence), microscopy (presence of acid-fast bacilli in pleural fluid/ pleural biopsy), culture (in pleural fluid/ pleural biopsy), histopathology (pleural biopsy showing the presence of typical caseating granulomatous inflammation), Xpert MTB/RIF or Xpert MTB/RIF Ultra positivity, elevated ADA (>40 IU/L) and response to ATT (improvement in effusion after starting ATT) was utilized for diagnosis of TBP patients. The patients showing positivity with any one of the tests from CRS were categorized as TBP. The patients presenting with pleural effusions showing the presence of cytological or histological evidence of malignant cells/etiology of other respiratory infections in pleural fluid/pleural biopsy but not showing the presence of *M.tb* in pleural fluid/biopsy were classified as non TB pleuritis (Non TBP) patients.

### Mycobacterial RNA isolation from pleural biopsy and pleural fluid

Pleural biopsy samples in RNA later were centrifuged at 3000g for 15 minutes at 4°C after adding 1 mL phosphate buffer saline (PBS) and was thereafter washed twice with PBS at 5000g for 15 minutes at 4°C. After washing, the biopsy samples were homogenised and resuspended in trizol. RNA isolation was then carried using trizol method as described by Abhishek et al, 2018 [27]. RNA was also isolated from pleural fluid samples after pelleting down the mycobacteria at at 4500g for 20 minutes at room temperature using same protocol. As positive control, RNA from log phase *M.tb* H37Rv culture grown in Soutan’s media using TRIzol as described by Abhishek et al [27]. The concentration and purity of isolated RNA was checked on nanodrop and RNA integrity of samples which were used for microarray was analysed using 2100 Bioanalyzer (Agilent).

### M. tb whole genome microarray and data analysis

The transcriptome analysis of M. tb in TBP biopsy samples (n=4)/ *M.tb* H37Rv was carried out using whole genome microarray with 200ng input RNA using Agilent’s microarray platform as described by Abhishek et al, 2018. Briefly the RNA was amplified and labeled with Cyanine 3-CTP (Cy3) followed by purification and quantitation of amplified cRNA. The samples having specific activity >15(pmol Cy3/μgcRNA), werefurther hybridized to custom designed microarray slides and scanned on the Agilent Sure Scan Microarray Scanner followed by image extraction and data analysis as described by Abhishek et al, 2018. Functional categorization of differentially expressed genes (DEGs) was performed according to TubercuList database. To delineate the metabolic pathways specifically expressed in TBP patients, data obtained from *M.tb* whole genome microarray was further analyzed by using BioCyc database.(BioCyc.org)

### qRT-PCR (Microarray validation)

The data attained from microarray was further confirmed by quantitative real time PCR. The Specific primers for *M.tb* genes were designed using Primer BLAST (supplementary table 1). Unamplified RNA (1µg-2µg) from biopsy samples of TBP patients and *in-vitro* grown *M.tb* H37Rv culture (Control) were DNase treated followed by cDNA synthesis using kit from Bio-Rad Laboratories as per manufacturer’s instructions and used for qRT-PCR on Qiagen Rotor-Gene Q realtime PCR machine using iTaq Universal SYBR Green Supermix® dye and *M.tb* gene-specific primers. For each gene, cDNA samples (reverse transcriptase positive, RT+) along with the no enzyme control (NEC, reverse transcriptase negative) were run in duplicates. No enzyme control reactions were used to check for DNA contamination. The amplification plot (CT, cycle threshold) and melt curve for the product specificity were analyzed. Comparative CT method (ΔΔCT) was used to determine relative gene expression of the target genes [27, 33] using 16srRNA as an endogenous control and *M.tb* RNA from *in-vitro* culture as a reference. Student’s t test with p<0.05 was used for statistical analysis.

### Development and validation of qRT-PCR assay for the diagnosis of tuberculous pleuritis

The mycobacterial transcriptomic signatures identified from pleural biopsy samples were further used for the development and validation of a mycobacterial RNA based molecular assay for the diagnosis of TBP using pleural fluid samples. A computer-generated random number table for total 216 TBP suspects included in this study was used and 1/4^th^ (n=53) of the study subjects with known clinical diagnosis (TBP and non TBP disease controls) were allocated to development cohort for the development of molecular assay. Remaining 3/4^th^ (n=163) of the study subjects were assigned to validation cohort for the validation of developed molecular assay using blinded samples with no information on clinical diagnosis at the time of assay. Two of the top upregulated mycobacterial genes in pleural biopsy samples as identified by microarray data and validated through qRT-PCR were used for the development and validation of RNA based diagnostic assay using pleural fluid samples. Briefly, isolated *M.tb* RNA from the pleural fluid of all the patients was assayed for presence or absence of two selected genes using qRT-PCR as described above.

The presence or absence of *M.tb* RNA in the pleural fluid samples was analyzed and a sample was considered as positive for TB when mycobacterial gene specific RNA was detected in duplicate tubes with Ct value (cycle threshold) less than 35 along with transcript specific melt peak. The RT+ sample was considered as positive for the presence of *M.tb* RNA only if the RT+ and RT-samples differed in ≥1 Ct. [27].

## RESULTS

In current study, total 261 TB pleuritis (TBP) suspects were recruited, out of which, 45 were excluded owing to indeterminate clinical diagnosis (n=24) or development of TB of any other site (n=7) or having both TB and malignancy simultaneously (n=4) or having other infectious diseases including HCV and COVID (n=4) or insufficient quantity of isolated RNA (n=6). Finally, 216 TBP suspects were included in the study. These study subjects (n=216) were divided into pleural TB (n=54) and non-TB controls (n=162) as per the composite reference standard (Figure1). Non-TB controls included patients with malignant pleural effusion (n=120) and other respiratory diseases (n=42) (Figure 1). Table 1 and 2 depict the demographic and clinical details of recruited study subjects.

**Figure 1:**
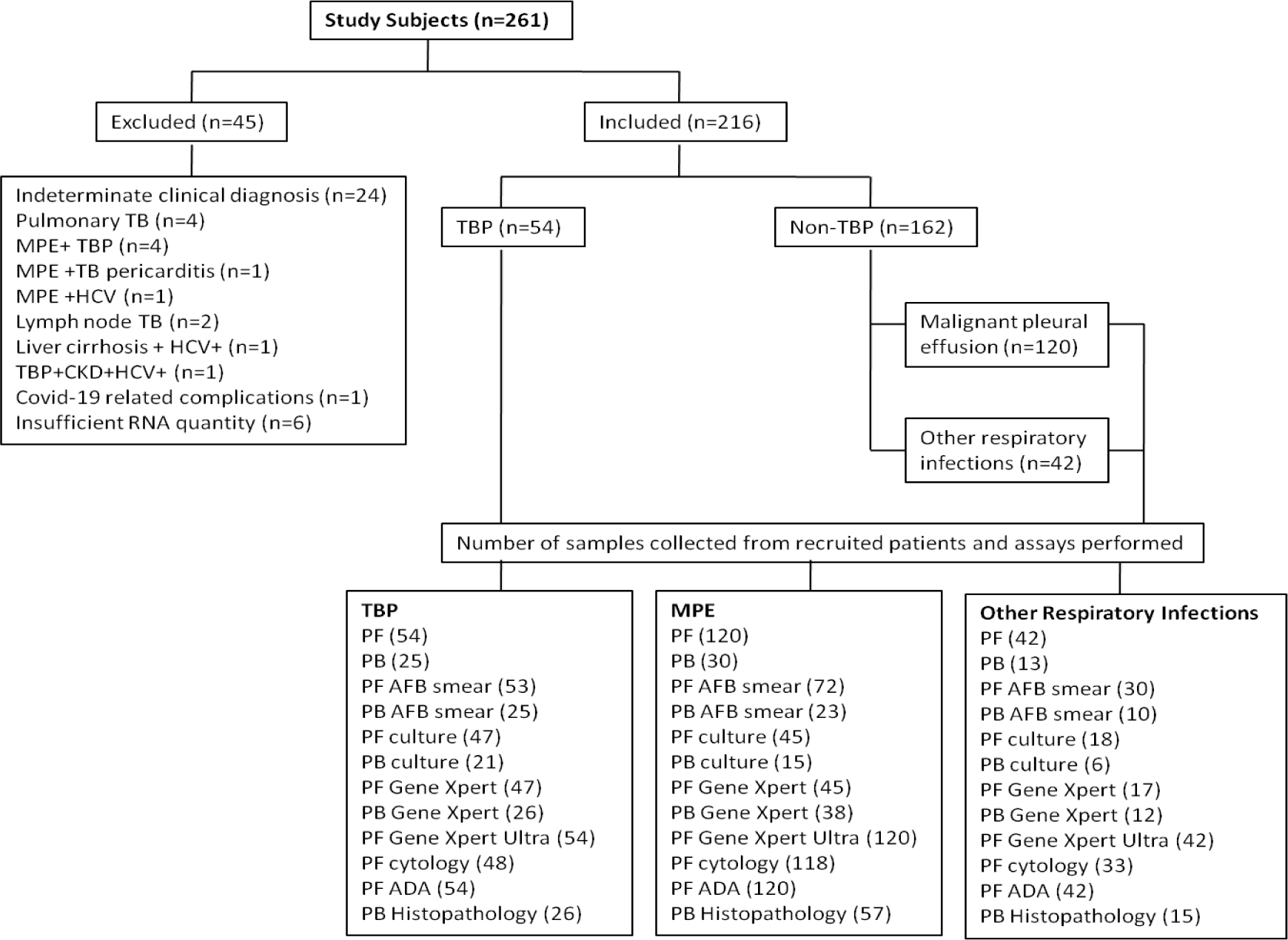
Consort diagram for the study showing details of enrolled study subjects. Values in parentheses show the number of patients in respective categories. ADA: Adenosine deaminase, AFB: Acid fast bacilli, PB: Pleural biopsy, PF: Pleural fluid, CKD: Chronic kidney disease, HCV: Hepatitis C virus, MPE: Malignant pleural effusion, TBP: TB pleuritis.

**Table 1:** Demographic profile of TBP and non TBP subjects.

| **Characteristics** | **TBP**<br>**(n=54)** | **Non-TBP**<br>**(n=162)** |
| --- | --- | --- |
| **Age (yr)** | 39.5 | 55.5 |
| **Males, n (%)** | 39 ( 72.22%) | 86 (53.08% ) |
| **HIV status** |  |  |
| **Positive** | 0 (0%) | 0 (0%) |
| **Negative** | 18 ( 33.33 %) | 33 (20.37%) |
| **Status Unknown** | 36 (66.66%) | 129 (79.62%) |
| **Mantoux Status** |  |  |
| **Positive** | 7 (12.96%) | 5 (3.08%) |
| **Negative** | 7 ( 12.96 %) | 11 (6.79%) |
| **Status Unknown** | 40 (74.07%) | 146 (90.12%) |
| **BCG status** |  |  |
| **Yes** | 45 (83.33%) | 143 (88.27 %) |
| **No** | 9 ( 16.66 %) | 11 (6.79%) |
| **Status Unknown** | 0 (0%) | 8 (4.93%) |

TBP: TB pleuritis, HIV: Human immunodeficiency syndrome, BCG: bacille Calmette-Guerin

**Table 2:**
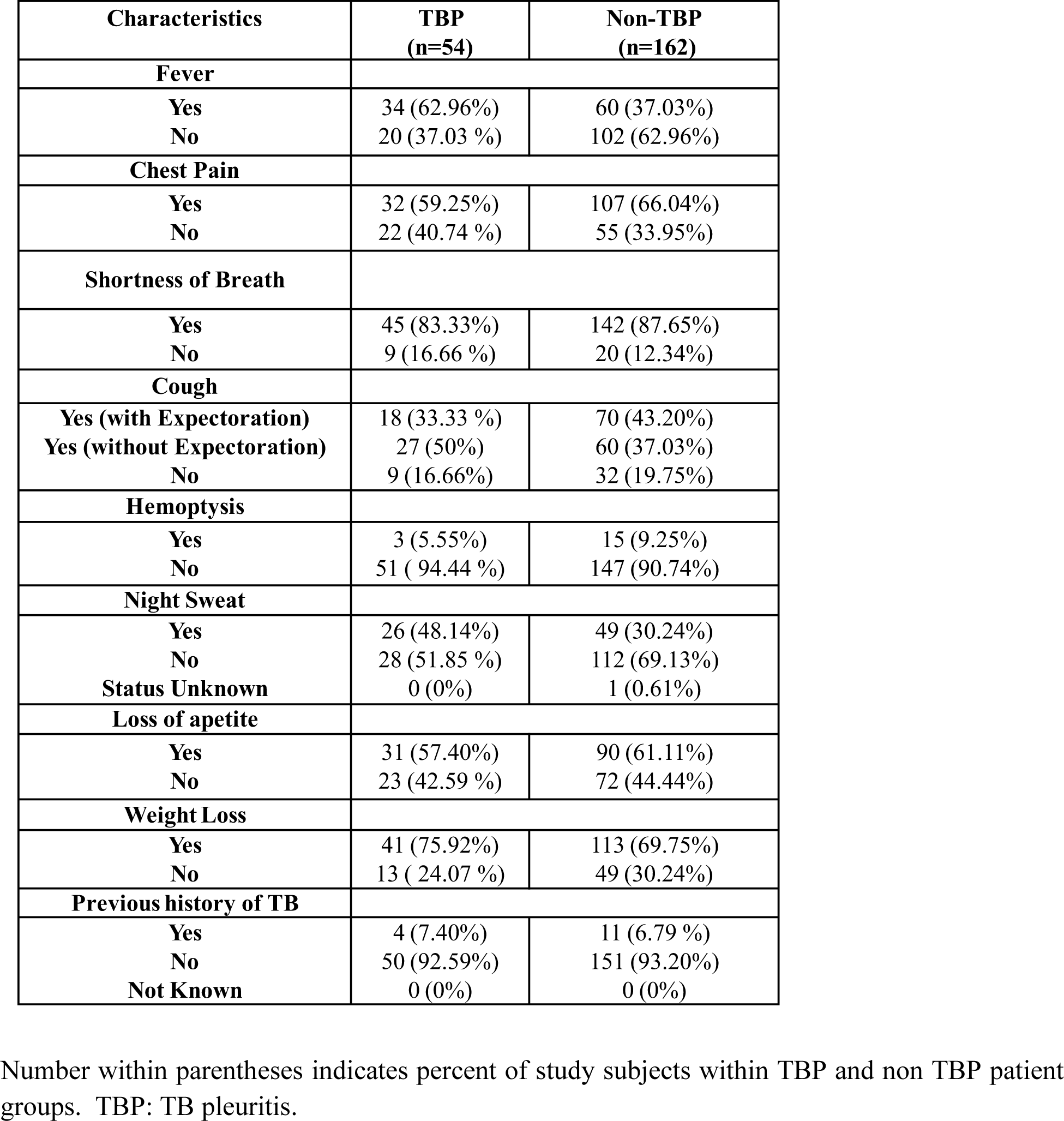
Clinical characteristics of TBP and non TBP study subjects.

### Transcriptomic profile of *Mycobacterium tuberculosis* (*M.tb*) in pleural biopsy samples of TBP patients

Whole genome microarray of *M.tb* was carried out in pleural biopsy samples from confirmed TBP patients (n=4) in comparison to in-vitro grown *M.tb* H37Rv culture. Overall whole genome microarray of mycobacteria in TBP patients led to identification of 1856 differentially expressed genes (DEGs) including 1365 upregulated and 491 downregulated genes (Supplementary table 3& 4). (Figure 2b & c) *The* differentially expressed mycobacterial transcripts were functionally categorized as per the Tuberculist database [34] (Figure 2d). Though most of the functional categories were enriched equally among the upregulated and downregulated genes, however, lipid metabolism and PE-PPE family genes were enriched only among upregulated genes (Table 3). Pathway analysis using BioCyc database [35] led to identification of significantly enriched (*p*<0.05) metabolic pathways among both the upregulated and downregulated categories. A significant enrichment among the upregulated genes was observed in pathways belonging to fatty acid, cholesterol and sugar/galactose degradation along with some other pathways as those belonging to pyrimidine biosynthesis, transcription regulation, DNA repair and metal ion transport (supplementary table 5). On the contrary a significant enrichment among downregulated genes was observed in pathways belonging to translation and protein secretion and localization. Genes belonging to stress responses like hypoxia, oxidative stress, nutrient stress, starvation and response to host defense pathways were also significantly enriched among the downregulated genes (supplementary table 6).

**Figure 2:**
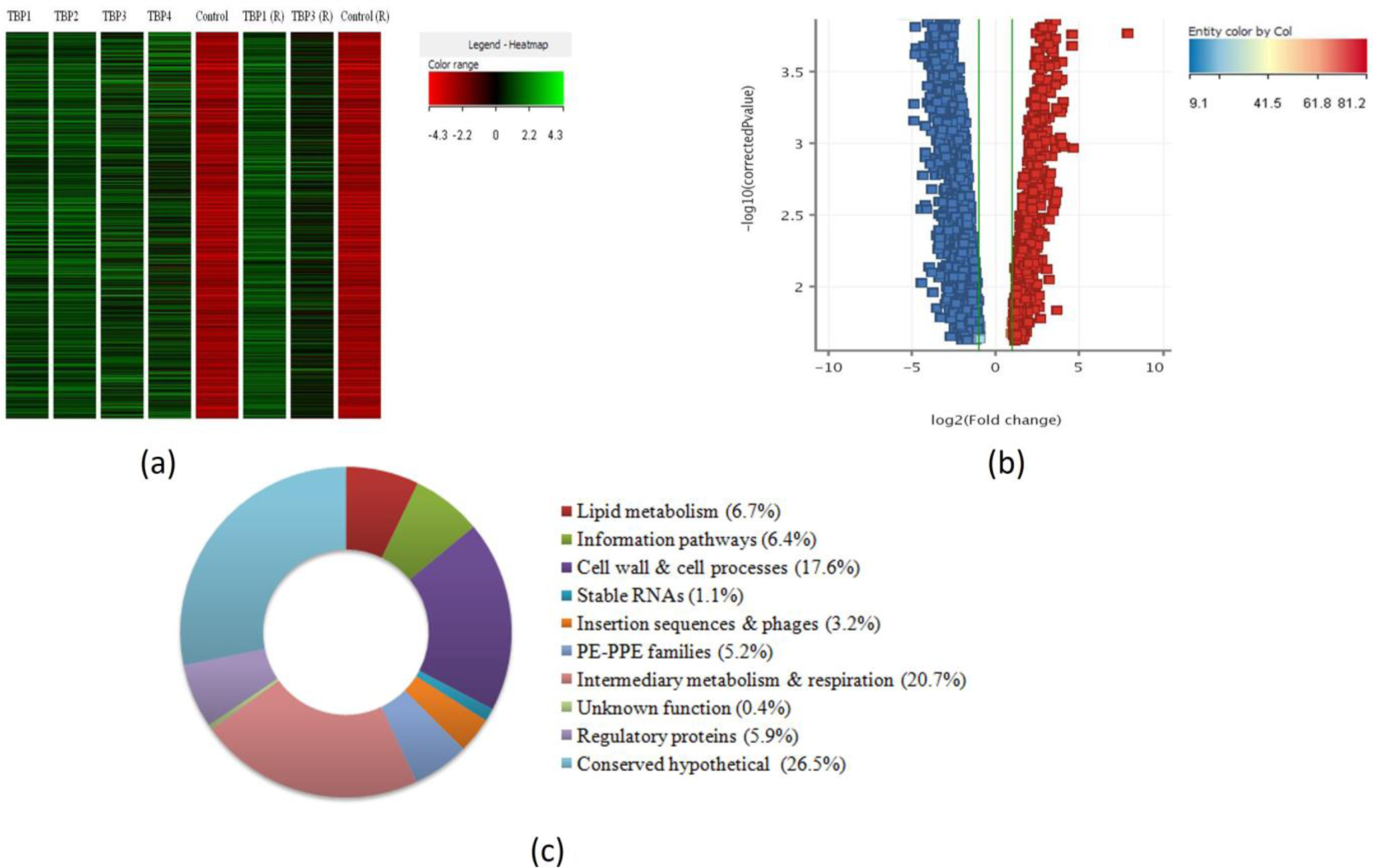
Transcriptome analysis of *M.tb* in biopsy samples of TBP patients through *M.tb* whole genome microarray. a) Heat map of mycobacterial transcriptome obtained in TBP and control samples (*in vitro* grown *M.tb* H37Rv culture) as obtained in whole genome microarray. Lane 1-4 (TBP samples), Lane 5 (control; *in vitro grown M.tb* H37Rv culture), lane 6 (technical replicate of TBP sample 1), lane 7 (technical replicate of TBP sample 3) and lane 8 (technical replicate of control sample). b) Volcano plot of differentially expressed (upregulated (red) and down regulated (blue)) mycobacterial genes in biopsy samples from TBP patients as analyzed by GeneSpring software. Log2 fold change is represented on x-axis and log10 adjusted p-values on Y-axis. *M.tb: Mycobacterium tuberculosis,* TBP: TB pleuritis, RIN: RNA integrity number. c) The pie chart representing the percentage of differentially expressed *M.tb* genes in each functional category as per the Tuberculist database identified in the biopsy samples of TBP patients by *M.tb* whole genome microarray. TBP: TB pleuritis *M.tb: Mycobacterium tuberculosis*

**Table 3:**
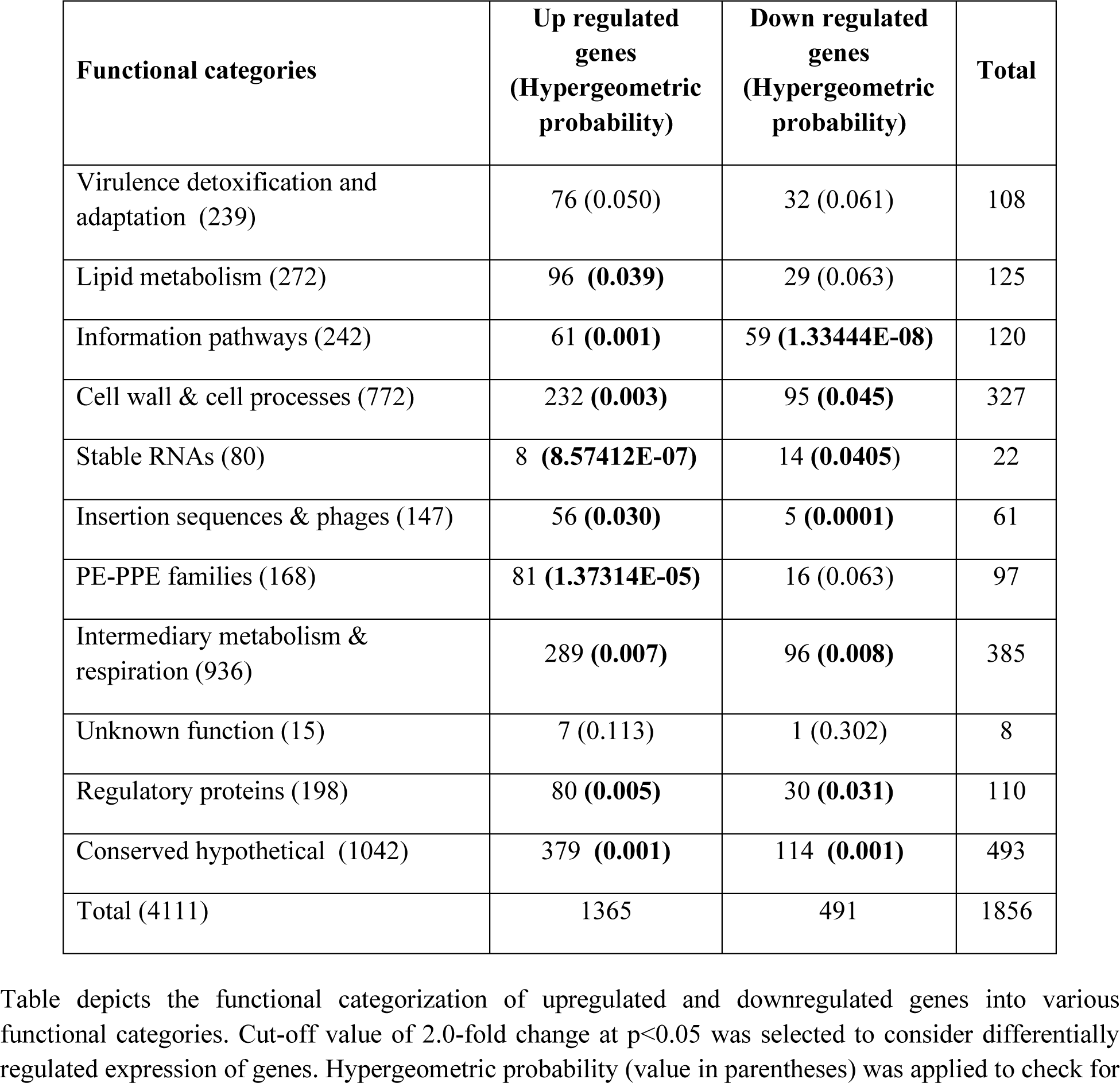
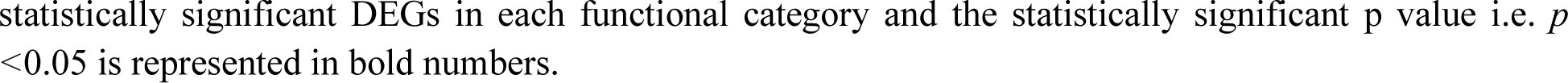
Functional categorization of differentially expressed mycobacterial genes (DEGs) in the biopsy samples of TBP patients.

| Functional categories | Up regulated genes<br>(Hypergeometric probability) | Down regulated genes<br>(Hypergeometric probability) | Total |
| --- | --- | --- | --- |
| Virulence detoxification and adaptation (239) | 76 (0.050) | 32 (0.061) | 108 |
| Lipid metabolism (272) | 96 (**0.039**) | 29 (0.063) | 125 |
| Information pathways (242) | 61 (**0.001**) | 59 (**1.33444E-08**) | 120 |
| Cell wall & cell processes (772) | 232 (**0.003**) | 95 (**0.045**) | 327 |
| Stable RNAs (80) | 8 (**8.57412E-07**) | 14 (**0.0405**) | 22 |
| Insertion sequences & phages (147) | 56 (**0.030**) | 5 (**0.0001**) | 61 |
| PE-PPE families (168) | 81 (**1.37314E-05**) | 16 (0.063) | 97 |
| Intermediary metabolism & respiration (936) | 289 (**0.007**) | 96 (**0.008**) | 385 |
| Unknown function (15) | 7 (0.113) | 1 (0.302) | 8 |
| Regulatory proteins (198) | 80 (**0.005**) | 30 (**0.031**) | 110 |
| Conserved hypothetical (1042) | 379 (**0.001**) | 114 (**0.001**) | 493 |
| Total (4111) | 1365 | 491 | 1856 |

Table depicts the functional categorization of upregulated and downregulated genes into various functional categories. Cut-off value of 2.0-fold change at $p < 0.05$ was selected to consider differentially regulated expression of genes. Hypergeometric probability (value in parentheses) was applied to check for
statistically significant DEGs in each functional category and the statistically significant p value i.e. $p < 0.05$ is represented in bold numbers.

### Microarray validation using qRT-PCR

Microarray data was validated using real time platform in biopsy samples with a subset of 10 genes including six upregulated genes (*Rv1586c, Rv2819c, Rv1971, Rv1582c, Rv2351c and Rv0974c)* and four downregulated genes (*Rv3872, Rv0983, Rv3875* and *Rv3871)* selected from the microarray analysis of TBP patients. Similar to the microarray data, the topmost upregulated genes *Rv1586c, Rv2819c* and *Rv1582c* showed an upregulated relative expression in real time RT-PCR with log2 fold change of 14.11,13.64 and 12.43 respectively (Figure 3). However, in case of *Rv1971* and *Rv0974*c, no amplification was observed in any of the tested samples whereas in case of *Rv2351c*, amplification was seen in only one (Log2FC 0.92) sample. The downregulated genes *Rv3872, Rv3875, Rv0983* selected from microarray data showed downregulation in biopsy samples of TBP patients by qRT-PCR also (Figure 3) thus validating the microarray results. In case of *Rv3871*, no amplification and melt curve were observed in samples. Hence, the results of microarray data could be validated for 6/10 selected genes (Figure 3).

**Figure 3:**
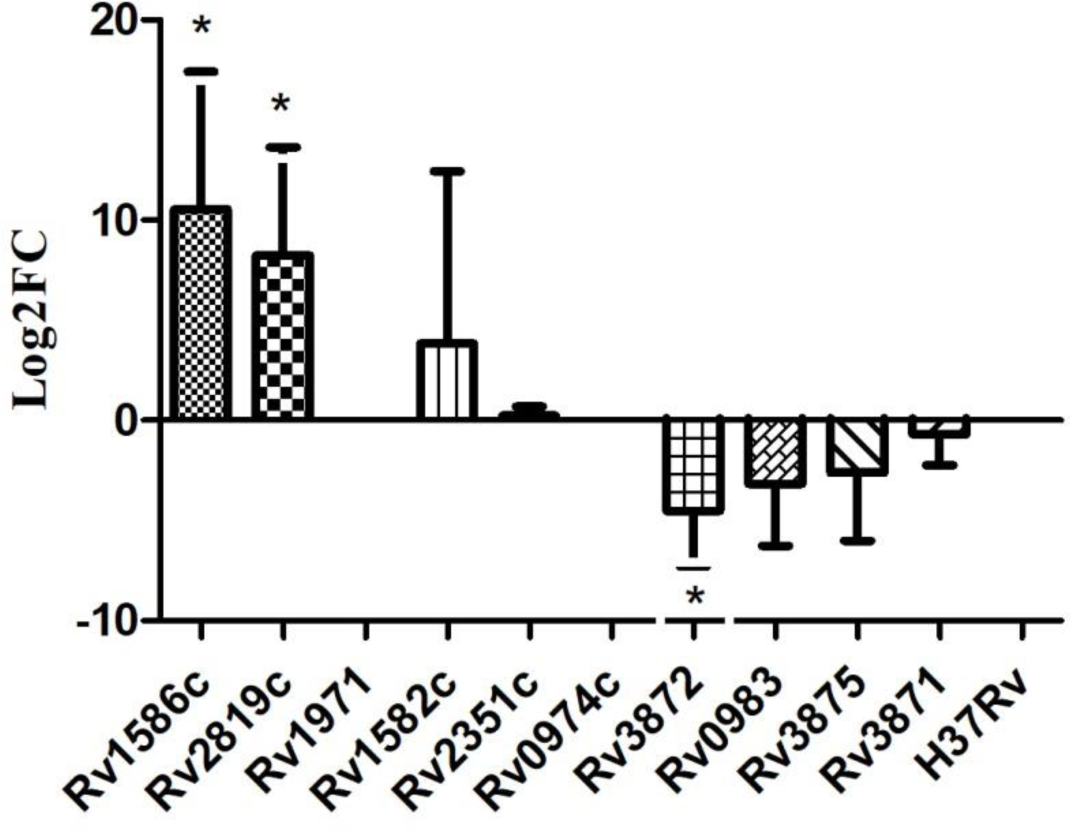
Validation of *M.tb* whole genome microarray by qRT-PCR. The relative expression of genes in both the upregulated (n=6) as well as downregulated (n=4) categories was analysed using RNA isolated from biopsy samples of TBP patients in comparison to *in-vitro* grown *M.tb* H37Rv. The bar graph represents log2 fold change in gene expression on y-axis, where positive values on y-axis indicate upregulation and negative values on y-axis indicate downregulation. For normalization, 16S rRNA was employed as reference gene(*p<0.05using student t-test for comparing the relative expression of each gene with *in-vitro* grown *M.tb* H37Rv). Each bar indicates log2 fold change in gene expression for the each of the gene obtained in 5-8 TBP samples with each sample analyzed in duplicates. *M.tb: Mycobacterium tuberculosis,* TBP: TB pleuritis. The relative expression of *Rv1586c* and *Rv2819c* was analyzed in 8 TBP biopsy samples and the expression of *Rv1582c* was analyzed in 5 TBP biopsy samples.

### Development and validation of RNA based molecular assay

#### Development of molecular assay for TB pleuritis

For developing real time RT-PCR based test using pleural fluid as sample, two topmost differentially upregulated *M.tb* genes *Rv1586c* (log2 FC: 9.89) and *Rv2819c* (FC: 8.09) whose expression were also validated through qRT-PCR were selected. To develop the assay, qualitative expression of selected genes i.e. *Rv1586c* and *Rv2819c* was first assessed in development cohort consisting of 10 TB pleuritis patients and 43 non-TB disease controls. Among the TBP patients, *Rv1586c* was detected in 6/10 patients thus showing sensitivity of 60%, however it also showed false detection in 3/43 disease controls thus giving a specificity of 93.02%. The other gene *Rv2819c* was detected in 7/10 TBP patients, showing a sensitivity of 70% and was falsely detected in 1/43 disease controls, thus having a specificity of 97.67 (Figure 4).

**Figure 4:**
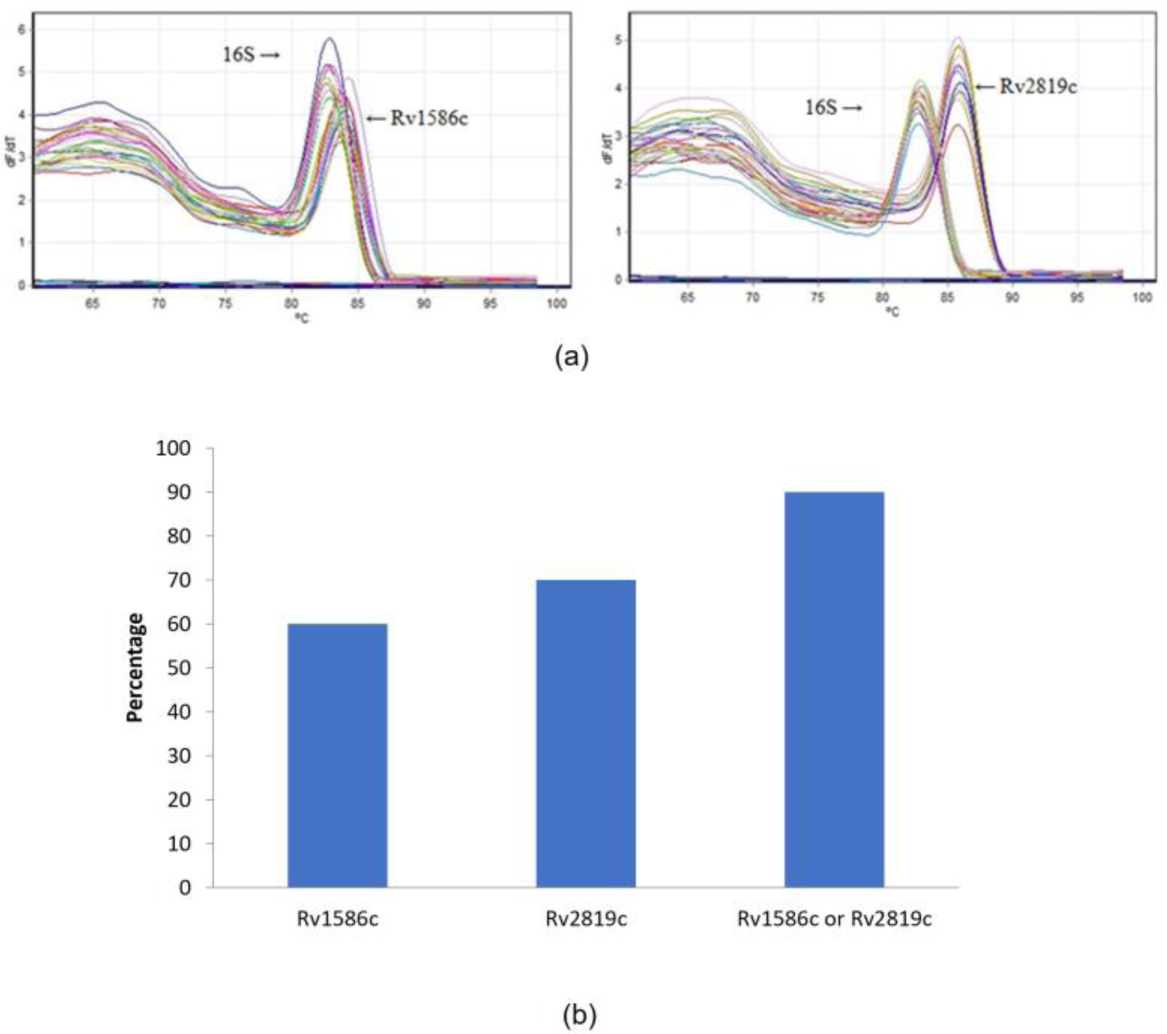
Development of qRT-PCR assay using mycobacterial genes *Rv1586c* and *Rv2819c* in pleural fluid samples of TBP and non-TBP subjects. a) Representative melt curve, b) The diagnostic performance of qRT-PCR. The data was analyzed by considering the positivity for *Rv1586c, Rv2819c* individually and either of the genes in each sample. Sample was considered positive based on Ct value (cut off 35) and presence of specific melt peaks for genes. Sensitivity=(TP)/(TP+FN), Specificity= (TN)/(FP+TN). Sen: Sensitivity, Spec: Specificity, TP: True positive, FN: False negative, TN: True negative, FP: False positive.

To increase the sensitivity of overall assay the data was further analyzed by looking at the presence of either of the genes for positive result. The analysis increased the sensitivity of detection to 90% with slight loss in the specificity which came out to be 93.02% (3/43). Thus, this analysis algorithm was taken forward for the evaluation of these genes in validation cohort (Figure 4b).

### Validation of assay

*Rv1586c and Rv2819c*based molecular assay was further validated in the blinded pleural fluid samples of validation cohort consisting of 163 TBP suspects. In the blinded analysis, using the algorithm from development cohort resulted in positivity in 43 samples with negative results in 120 samples (supplementary Figure 1).

After decoding, the true categorization of patients based on their final diagnosis using various laboratory and clinical criteria as described in methodology (Annexure III). Of 44 BP subjects, 35 samples were found as true positive and 9 subjects were falsely detected as negative by Amongst 119 non TBP subject, 111 samples were identified as negative and 8 were detected as falsely positive by qRT-PCR. Hence based on these results, the real time RT-PCR assay had a sensitivity of 79.55% (35/44 TBP) with specificity of 93.28% (8/119 non TBP patients) (Figure 5). Further on comparative evaluation of diagnostic potential of qRT-PCR based on these selected genes with available diagnostic modalities for TBP, real time RT-PCR assay showed better performance than either of existing diagnostic tests in pleural fluid (Supplementary table 8) except for histopathology that showed highest sensitivity and specificity of 94.74% and 100% respectively.

**Figure 5:**
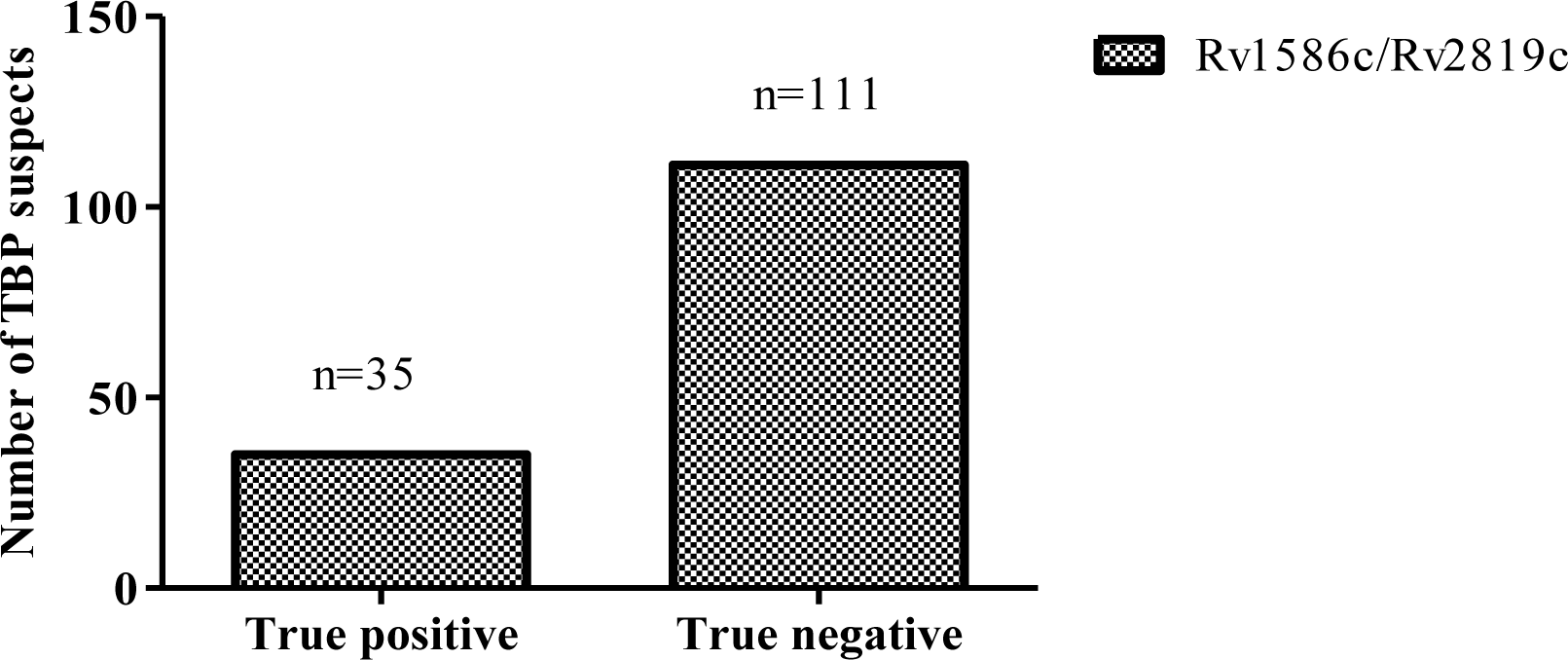
Performance of developed real time RT-PCR assay in pleural fluid samples of validation cohort. TBP suspects (n=163) were assessed for the presence of either of mycobacterial genes *Rv1586c/Rv2819c* and data was compared with the final diagnosis obtained for each patient using composite reference standard to calculate the number of True positive (n=35), True Negative (n=111), False Positive (n=8) and False Negative (n=9) patients.

## DISCUSSION

Routine diagnostic techniques utilizing pleural fluid suffer from limitations and do not provide reliable performance for TBP diagnosis. We hypothesized that the biomarkers which are expressed *in-vivo* at the site of infection during disease condition will serve as important diagnostic targets. Therefore, the mycobacterial transcriptome was studied in the biopsy samples obtained from confirmed TBP patients considering the fact that mycobacterial RNA isolated from pleural fluid might not be of optimum quantity and quality for microarray. Transcriptomic profile of *M.tb* in the biopsy samples from TBP patients demonstrated 1856 differentially expressed genes (DEGs) as compared to *in-vitro* grown *M.tb* H37Rv. The number of upregulated genes was higher in comparison to downregulated genes, a pattern similar to that observed in M*.tb* infected alveolar epithelial cells and macrophages but contrary to sputum samples of PTB patients [25,28,36,37]. The *in-vivo* upregulated mycobacterial transcripts in pleural biopsy samples were further used for the development of RNA based molecular assay in pleural fluid samples.

Several studies have previously used host transcriptomic signatures for TB diagnosis [38–40]. However, pathogen specific transcriptomic signatures seem to be more important for the specificity of an assay. For exploring the role of mycobacterial transcripts for the development of a molecular assay for pleural TB, microarray results were first validated on real time quantitative reverse transcription PCR (qRT-PCR) using unamplified RNA isolated from pleural biopsy samples obtained from TBP patients. Six out of 10 genes assessed by qRT-PCR showed same expression pattern as that observed in microarray. Some level of disagreement between microarray and qRT-PCR has also been observed in previous studies [26, 29] which may be due to multiple factors like the handling and storage conditions of samples, differences in the data interpretation strategies etc. [41]We selected two of the top upregulated genes validated through qRT-PCR i.e., *Rv1586 and Rv2819c* belonging to different functional categories for development of an RNA based molecular assay using the pleural fluid samples to discriminate TBP and non TBP patients. *Rv1586c* (Probable PhiRv1 integrase) belongs to the category of Probable PhiRv1 phage proteins of prophage-like elements ϕ Rv1 of *M.tb* and encodes putative integrase of the serine recombinase type. The prophage-like elements ϕ Rv1 is specifically expressed in *M.tb* and absent in BCG. The second selected gene Rv2819c, encodes for a hypothetical protein (375 amino acid) and not much is known about it.

The qRT-PCR assay based on these two genes *Rv1586c* and *Rv2819c* was initially developed in study subjects comprising development cohort and having known diagnosis on the basis of routine laboratory and clinical diagnostics, however blinded samples were used for validation part of the study. The data of qRT-PCR was analysed using the MIQE guidelines which have previously been used by several studies reporting qRT-PCR data including the diagnostic assays developed for COVID testing based on qRT-PCR [42–44]. Using these criteria, when positivity of only *Rv1586c* or *Rv2819c* was considered, it led to diagnosis of TBP patients with sensitivity and specificity of 60-70% and 93-97% respectively. Analysis of data indicated that although many of the samples showed positivity for both the genes, but several samples also showed the positivity to either of these two genes. Hence, to further improve the sensitivity of the assay, positivity of either of genes for a positive qRT-PCR test was considered which resulted in improved sensitivity without significant compromise in specificity for differentiating TBP patients from non TBP study subjects. Hence for validation cohort, results were analyzed considering the positivity for either of the genes (*Rv1586c* / *Rv2819c*) in the same pleural fluid sample. Decoding the qRT-PCR results obtained in TBP suspects of validation cohort based on the CRS indicated 79.6% sensitivity and 93.02% specificity of the assay for TBP diagnosis. In comparison to the RNA based assay developed in this study, the sensitivity of Xpert MTB/RIF and Xpert MTB/RIF Ultra was lower (Supplementary Table 6). Previous studies have also shown low sensitivity of Xpert MTB/RIF in pleural fluid [45–47]. although recent studies on Xpert MTB/RIF Ultra have shown improved performance over Xpert MTB/RIF [24,48,49]. The test developed in the current study although has a better sensitivity than ultra, however specificity is slightly lower. Moreover, the test being an RNA based assay could be advantageous over DNA based Xpert MTB/RIF and Ultra that cannot discriminate between live and dead bacilli leading to false positivity in previously treated individuals [25, 50]. But assay developed in current study cannot be used for detection of antibiotic resistance as in case of Xpert MTB/RIF and Xpert MTB/RIF Ultra. Though RNA based assay will not be a user-friendly assay and will require trained personnel, but the widespread use of a similar assay for diagnosing SARS-COV2 during the COVID pandemic has provided mankind with enough experience to handle such assays even at hospitals other than tertiary care centers.

As a secondary objective, the in vivo expressed mycobacterial transcriptome data was also analyzed in depth to understand the pathogenesis of pleural TB. The differentially expressed upregulated and downregulated genes were categorized into different functional categories as per tuberculist database [34]. Genes of PE-PPE family and lipid metabolism were significantly enriched in the upregulated pathways (Table 3). Number of PE-PPE family proteins are implicated in M.tb pathogenesis via promoting the attachment of microbe to the host cells [51,52]. A significant enrichment of fatty acid degradation, utilization and assimilation along with cholesterol degradation pathways was observed among the upregulated category. Fatty acids and lipids including cholesterol are vital components of mycobacterial cell wall and also essential for carbon and energy sources for *M.tb* adaptation in host microenvironment to establish and maintain chronic TB infection [53–57]. Pyrimidine biosynthesis, transcription regulation and DNA repair pathways known to play an important role during *M.tb* infection and disease progression were also upregulated [58, 59]. Several down regulated pathways were also significantly enriched and primarily included translation, protein secretion and localization similar to that reported previously in sputum mycobacterial cells [26,37] and could be considered as an adaptive strategy in host environment [60]. Overall, it seems that mycobacteria in pleural space is engaged in energy generation using fatty acid degradation and is increasing its virulent properties with the upregulation of PE-PPE genes. These genes along with DNA repair genes are also helping the bacteria to deal with the host immune system and various stresses.

In conclusion, the transcriptome analysis of *Mycobacterium tuberculosis* in biopsy samples of TB pleuritis patients led to newer insight into the in-vivo expression of mycobacterial transcriptomic signatures in the pleural space during TBP. The selected pathogenic RNA signatures from the M. tb transcriptome in TBP which are highly expressed in-vivo have shown promising results for the development of RNA based diagnostic real time PCR assay using pleural fluid. However, it needs further optimization to develop it into user friendly assay devoid of technical complexities of real time RT-PCR.

## Data Availability

The gene expression data obtained in the current study has been submitted to Gene Expression Omnibus (GEO) vide accession no. GSE231472.

