## Supplementary Table/ Figure for "Diagnostic utility of in vivo expressed mycobacterial RNA transcripts in pleural fluid for the differential diagnosis of tuberculous pleuritis"

| S. No. | Gene Name | Primer Sequence (5'<-----Sequence----->3') |
| --- | --- | --- |
| 1 | Rv1586c | Forward primer: GACAGCTCGGCTATCATCCC  Reverse primer: CAACAGGATCAGGAACGGCT |
| 2 | Rv2819c | Forward primer: TGAAAGCGTGAATCTCGTTG  Reverse primer: AAGAGTGGGTAGAGCCAAAC |
| 3 | Rv2351c | Forward primer: TCCCTGCATTGTCATTTCGC  Reverse primer: TGGAGTCGCAAAGTTGAACG |
| 4 | Rv1971 | Forward primer: CTCACCTATCACAACGACATCG  Reverse primer: CCTGCTTGGTGTTGAGGTTG |
| 5 | Rv1582c | Forward primer: CTACTTCAAACGGCACCACT  Reverse primer: CATACCCCACTGCTCATCAC |
| 6 | Rv0974c | Forward primer: ATGCTTGATCATCCCGCC  Reverse primer: CGCCGCTGTTGTTCTTAC |
| 7 | 16S | Forward primer: TTGTCTCATGTTGCCAGCAC  Reverse primer: ACCGGCTTTTAAGGATTCGC |
| 8 | Rv3872 | Forward primer: TTCACATCGGAGGGCATC  Reverse primer: TCGTCGATTTGCGAATAGGT |
| 9 | Rv0983 | Forward primer: CATGACGAATCACCCACG  Reverse primer: CAGTCGAACTGCTGGCTG |
| 10 | Rv3875 | Forward primer: TCCATTCATTCCCTCCTTGA  Reverse primer: TTTGCTGGACACCCTGGTA |
| 11 | Rv3871 | Forward primer: ACCTGTCAGATGAGCCAGGC  Reverse primer: TTCTTCTGGAGGCTCGATGT |

**Supplementary table 1: Primer sequences of selected mycobacterial genes for real time**

**Supplementary table 2 and 3 are attached separately.**

**Supplementary table 4: List of significantly enriched (p<0.05) upregulated mycobacterial pathways identified in biopsy samples of TB pleuritis patients using BioCyc database.**

| **S.No** | **Pathways** | **p-values** | **No. of genes** | **Gene ID** |
| --- | --- | --- | --- | --- |
| 1 | Fatty Acid and Lipid Degradation | 0.004141007 | 54 | fadB3,Rv1136,echA9,echA18,echA17,echA14,fadE32,fadA6,ipdA,ipdB,ipdC,fadE30,hsaG,Rv1135A,echA13,echA15,echA7,echA16,echA18.1,fadE35,fadE36,fadE33,fadE31,fadE22,fadE17,fadE18,fadE9,fadE3,fadE1,fadD18,fadD17,fadD35,fadD1,fadD11,fadD11.1,fadD16,fadD8,fadD4,fadD7,fadD34,ltp2,Rv3541c, Rv3542c, fadE28, fadA5,fadE34,echA19,fadE26,  fadE27,glpD1, plcA, plcC, lipU, lipY |
| 2 | Cholesterol Degradation | 0.01984 | 22 | ltp2,Rv3541c,Rv3542c,fadE28,fadA5,fadE34,fadB3,echA19,fadE26,fadE27,Rv1136,echA9,echA18,echA17,echA14,fadE32,fadA6,ipdA,ipdB,ipdC,fadE30,hsaG |
| 3 | Sugar Degradation | 0.048174635 | 8 | hsaG,galE1,galE3,galTb,galTa,galK,xylB,Rv0648 |
| 4 | D-galactose degradation (galactose catabolic process via UDP-galactose) | 0.040384337 | 5 | galE1,galE3,galTb,galTa,galK |
| 5 | pyrimidine ribonucleoside monophosphate biosynthetic process | 0.002095334 | 7 | Upp,pyrF,pyre E,pyrC,pyrB,carB,carA |
| 6 | Transcription regulation | 0.013715146 | 99 | vapB23,Rv0775,Rv3405c,Rv1930c,relK,Rv0195,mce1R,sigC,virS,Rv3066,Rv0880,Rv3912,prpR,aldR,Rv3095,whiB5,vapB3,Rv2160A,Rv3575c,Rv0144,sigL,Rv1395,vapB12,nmtR,Rv1674c,Rv2250c,embR,Rv1219c,Rv2506,relG,Rv3557c,Rv1049,Rv1358,Rv2989,Rv0576,mazE9,Rv0339c,Rv1931c,Rv2640c,vapB19,nadR,Rv0196,sigG,Rv2327,Rv1725c,vapB9,Rv0767c,rslA,sufR,Rv0067c,Rv3413c,Rv2324,Rv0330c,cmr,ethR,Rv3055,vapB20,Rv2488c,Rv0792c,Rv0377,Rv2011c,Rv0324,Rv1685c,Rv0260c,Rv1776c,Rv0104,Rv0302,mazE3,espR,Rv0238,vapB7,vapB43,Rv1353c,sigM,relJ,tcrA,Rv0737,Rv0691c,Rv0494,Rv1773c,oxyS,Rv2282c,Rv3160c,vapB41,Rv2884,sigI,Rv0158,rskA,mazE7,Rv0653c,vapB14,Rv0328,Rv0825c,Rv3183,Rv0078,fmu,mce3R,Rv2642,Rv0918 |
| 7 | DNA repair | 0.000864416 | 34 | ruvA,recN,recD,recB,ruvC,Rv3201c,Rv3202c,radA,mutY,Rv0944,Rv2464c,nei,fpg,Rv3649,priA,ung,mpg,udgB,dnaE2,dinX,ligC,Rv3204,Rv3394c,Rv2415c,Rv1151c,ruvB,mutT3,mutT1,Rv3395c,ligA,ligB,Rv1000c,Rv3113,Rv1277 |
| 8 | Metal ion transport | 0.007503876 | 14 | corA,ctpD,mntH,Rv2025c,ctpG,Rv0318c,ctpC,irtB,irtA,dppA,dppB,dppC,Rv0203,ctpV |

**Supplementary table 5: List of significantly enriched (p<0.05) downregulated mycobacterial pathways identified in biopsy samples of TB pleuritis patients using BioCyc database.**

| **S.No.** | **Pathways** | **p-values** | **No. of genes** | **Gene ID** |
| --- | --- | --- | --- | --- |
| 1 | Translation | 0.00000009245184 | 36 | alas,frr,fusA1,infA,infC,rimP,rplB,rplE,rplF,rplI,rplK,rplM,rplP,rplQ,rplS,rplT,rplU,rplV,rplX,rpmA,rpmC,rpmD,rpmE,rpmG2,rpmH,rpmJ,rpsG,rpsJ,rpsL,rpsM,rpsN1,rpsO,rpsP,rpsR1,Rv2477c,tuf |
| 2 | Protein secretion and localization to extracellular region | 0.0000026000323 | 12 | eccB1,eccCa1,eccCb1,espA,espB,espC,espD,esxA,esxB,secE1,secG,tatA |
| 3 | Response to hypoxia | 0.00006741014 | 22 | Acg,ald,bfrB,cmaA2,devR,devS,fdxA,groEL2,hrp1,hspX,narX,Rv0569,Rv2005c,Rv2629,Rv3134c,Rv3371,sigB,TB31.7,tgs1,tgs2,trxB2,tuf |
| 4 | Response to oxidative stress | 0.007325844 | 16 | ahpC,bfrB,clgR,fhaB,furA,htpG,katG,lsr2,mpt53,rshA,Rv2455c,sigF,sigh,soda,tpx,trxC |
| 5 | Response to nutrients levels | 0.012334948 | 13 | bkdB,eccA3,grpE,hspX,hupB,mpt64,pckA,pstS1,relA,Rv2557,Rv2558,Rv3583c,sigF |
| 6 | Response to starvation | 0.02392517 | 12 | pckA,hupB,eccA3,relA,Rv3583c,sigF,pstS1,mpt64,Rv2558,Rv2557,hspX,grpE |
| 7 | Respinse to host defense pathways | 0.018851994 | 28 | Acg,clgR,cmaA2,desA1,eccB1,eccCa1,eccCb1,eccD1,espC,espH,espL,espR,esxA,fadD26,fadD28,fdxA,glnA1,hrp1,hspX,lpqH,mce1A,moeB1,pckA,prcB,Rv1462,Rv1738,Rv2336,secA2 |

**Supplementary table 6: Comparison of performance of the mycobacterial RNA based real time RT-PCR assay developed in the present study (based on the detection of either of mycobacterial genes *Rv1586c/Rv2819c)* in pleural fluid samples of TB pleuritis suspects with available diagnostic tests for TB pleuritis.PF: Pleural fluid, ADA: Adenosine deaminase, n in the parentheses indicate the number of patients in which test was performed.**

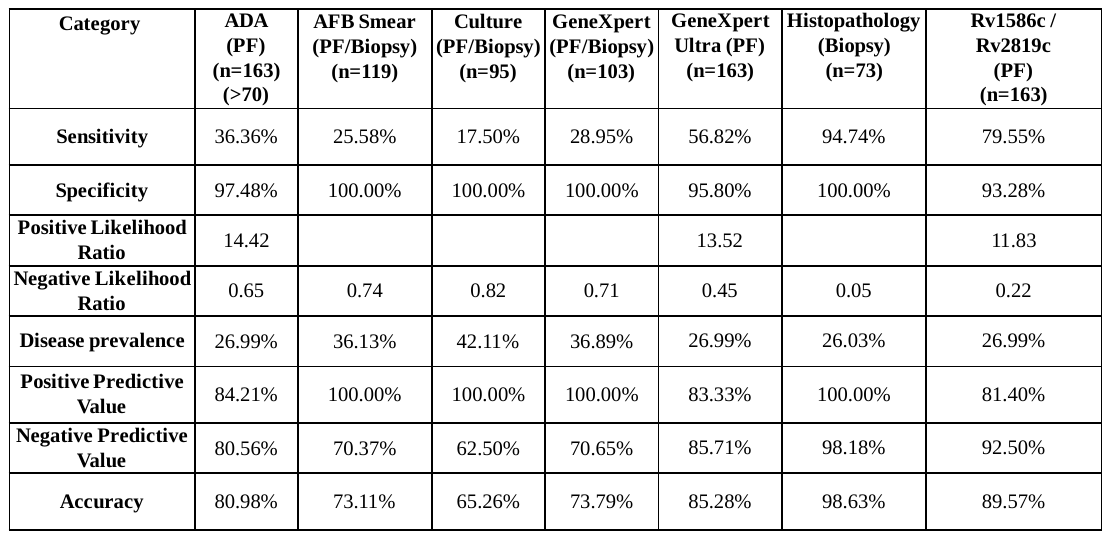

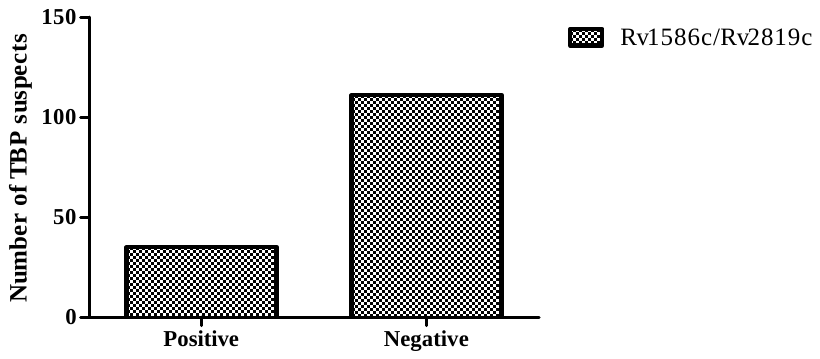

**Supplementary Figure 1: The results obtained in qRT-PCR based on detection of either of the mycobacterial genes (*Rv1586c/Rv2819c*) in blinded pleural fluid sample of study subjects from validation cohort.** The data was analyzed by considering the positivity for either of two genes in each sample. Sample was considered positive based on Ct value (cut off 35) and presence of specific melt peaks for genes. Samples showing no amplification /nonspecific melt peak for genes were considered negative.
