## Supplementary Table 2 for "Diagnostic utility of in vivo expressed mycobacterial RNA transcripts in pleural fluid for the differential diagnosis of tuberculous pleuritis"

**Supplementary Table 2: Upregulated genes in biopsy samples of tb pleuritis patients**

| **S.No** | **Gene Name** | **Gene ID** | **FC ([test] Vs [control])** |
| --- | --- | --- | --- |
| 1 | Rv1586c | Rv1586c | 949.73254 |
| 2 | Rv1582c | Rv1582c | 386.94852 |
| 3 | Rv2819c | Rv2819c | 272.42404 |
| 4 | Rv1576c | Rv1576c | 253.0093 |
| 5 | Rv0072 | Rv0072 | 234.06364 |
| 6 | Rv3513c | fadD18 | 233.06926 |
| 7 | Rv2817c | Rv2817c | 230.1762 |
| 8 | Rv2816c | Rv2816c | 223.75397 |
| 9 | Rv1762c | Rv1762c | 206.56569 |
| 10 | Rv0073 | Rv0073 | 203.30266 |
| 11 | Rv1587c | Rv1587c | 202.18518 |
| 12 | Rv2818c | Rv2818c | 198.11545 |
| 13 | Rv2543 | lppA | 190.13112 |
| 14 | Rv1578c | Rv1578c | 174.3045 |
| 15 | Rv1584c | Rv1584c | 164.88501 |
| 16 | Rv2544 | lppB | 156.9691 |
| 17 | Rv1579c | Rv1579c | 149.42424 |
| 18 | Rv1574 | Rv1574 | 133.25433 |
| 19 | Rv1761c | Rv1761c | 93.795944 |
| 20 | Rv1583c | Rv1583c | 78.79622 |
| 21 | Rv1575 | Rv1575 | 78.17494 |
| 22 | Rv1573 | Rv1573 | 67.71976 |
| 23 | Rv2263 | Rv2263 | 62.21458 |
| 24 | Rv1758 | cut1 | 52.12306 |
| 25 | Rv0978c | PE_PGRS17 | 46.3955 |
| 26 | Rv1759c | wag22 | 43.89951 |
| 27 | Rv1580c | Rv1580c | 42.504227 |
| 28 | Rv0575c | Rv0575c | 41.945393 |
| 29 | Rv1405c | Rv1405c | 41.84597 |
| 30 | Rv3352c | Rv3352c | 40.670303 |
| 31 | Rv1585c | Rv1585c | 39.59564 |
| 32 | Rv0953c | Rv0953c | 31.780813 |
| 33 | Rv3637 | Rv3637 | 31.343756 |
| 34 | Rv1971 | mce3F | 30.570276 |
| 35 | Rv0918 | Rv0918 | 30.205133 |
| 36 | Rv2191 | Rv2191 | 29.766378 |
| 37 | Rv2614A | Rv2614A | 28.839746 |
| 38 | Rv2736c | recX | 28.830454 |
| 39 | Rv2378c | mbtG | 28.178532 |
| 40 | Rv1135A | Rv1135A | 27.729437 |
| 41 | Rv0339c | Rv0339c | 26.68169 |
| 42 | Rv0739 | Rv0739 | 26.57113 |
| 43 | Rv2014 | Rv2014 | 25.931082 |
| 44 | Rv2886c | Rv2886c | 24.810728 |
| 45 | Rv0022c | whiB5 | 24.09142 |
| 46 | Rv1452c | PE_PGRS28 | 23.967117 |
| 47 | Rv3639c | Rv3639c | 23.835245 |
| 48 | Rv0194 | Rv0194 | 22.750341 |
| 49 | Rv2658c | Rv2658c | 21.857798 |
| 50 | Rv3539 | PPE63 | 21.817436 |
| 51 | Rv2262c | Rv2262c | 21.527384 |
| 52 | Rv0689c | Rv0689c | 21.288685 |
| 53 | Rv0979c | Rv0979c | 21.142756 |
| 54 | Rv2505c | fadD35 | 20.9768 |
| 55 | Rv3746c | PE34 | 20.514648 |
| 56 | Rv0895 | Rv0895 | 20.441334 |
| 57 | Rv2013 | Rv2013 | 20.069025 |
| 58 | Rv2249c | glpD1 | 19.834345 |
| 59 | Rv2363 | amiA2 | 19.282183 |
| 60 | Rv3345c | PE_PGRS50 | 19.064013 |
| 61 | Rv2998 | Rv2998 | 18.84673 |
| 62 | Rv2310 | Rv2310 | 18.81122 |
| 63 | Rv1290c | Rv1290c | 18.598759 |
| 64 | Rv1105 | Rv1105 | 18.594685 |
| 65 | Rv2667 | clpC2 | 18.567669 |
| 66 | Rv1541c | lprI | 18.395294 |
| 67 | Rv1572c | Rv1572c | 18.341831 |
| 68 | Rv1000c | Rv1000c | 18.319935 |
| 69 | Rv0661c | vapC7 | 18.242311 |
| 70 | Rv3195 | Rv3195 | 17.930506 |
| 71 | Rv3772 | hisC2 | 17.8904 |
| 72 | Rv2635 | Rv2635 | 17.88817 |
| 73 | Rv3834c | serS | 17.64117 |
| 74 | Rv0395 | Rv0395 | 17.058937 |
| 75 | Rv2891 | Rv2891 | 16.967539 |
| 76 | Rv3551 | Rv3551 | 16.924444 |
| 77 | Rv3517 | Rv3517 | 16.700153 |
| 78 | Rv1276c | Rv1276c | 16.619448 |
| 79 | Rv0603 | Rv0603 | 16.469612 |
| 80 | Rv0847 | lpqS | 16.241207 |
| 81 | Rv3638 | Rv3638 | 16.097675 |
| 82 | Rv3516 | echA19 | 16.081472 |
| 83 | Rv1268c | Rv1268c | 16.07737 |
| 84 | Rv3488 | Rv3488 | 16.032536 |
| 85 | Rv0191 | Rv0191 | 16.02781 |
| 86 | Rv1705c | PPE22 | 15.9283695 |
| 87 | Rv0393 | Rv0393 | 15.856407 |
| 88 | Rv3449 | mycP4 | 15.794931 |
| 89 | Rv0584 | Rv0584 | 15.468685 |
| 90 | Rv2384 | mbtA | 15.333814 |
| 91 | Rv0182c | sigG | 15.205804 |
| 92 | Rv2283 | Rv2283 | 15.129351 |
| 93 | Rv3912 | Rv3912 | 15.069107 |
| 94 | Rv3084 | lipR | 15.00384 |
| 95 | Rv1380 | pyrB | 14.984597 |
| 96 | Rv0372c | Rv0372c | 14.958994 |
| 97 | Rv0728c | serA2 | 14.897875 |
| 98 | Rv3839 | Rv3839 | 14.80448 |
| 99 | Rv1034c | Rv1034c | 14.734514 |
| 100 | Rv1933c | fadE18 | 14.692265 |
| 101 | Rv0654 | Rv0654 | 14.615826 |
| 102 | Rv3659c | Rv3659c | 14.606273 |
| 103 | Rv2807 | Rv2807 | 14.588455 |
| 104 | Rv1089A | celA2a | 14.575662 |
| 105 | Rv2550c | vapB20 | 14.574985 |
| 106 | Rv2593c | ruvA | 14.440174 |
| 107 | Rv2033c | Rv2033c | 14.201369 |
| 108 | Rv3776 | Rv3776 | 14.097024 |
| 109 | Rv2529 | Rv2529 | 14.052277 |
| 110 | Rv1581c | Rv1581c | 13.93191 |
| 111 | Rv1964 | yrbE3A | 13.903545 |
| 112 | Rv2671 | ribD | 13.902776 |
| 113 | Rv0600c | Rv0600c | 13.855483 |
| 114 | Rv1460 | Rv1460 | 13.752295 |
| 115 | Rv0618 | galTa | 13.745465 |
| 116 | Rv1938 | ephB | 13.711361 |
| 117 | Rv0614 | Rv0614 | 13.603106 |
| 118 | Rv1970 | lprM | 13.601141 |
| 119 | Rv2087 | Rv2087 | 13.590246 |
| 120 | Rv2265 | Rv2265 | 13.55327 |
| 121 | Rv3552 | Rv3552 | 13.53735 |
| 122 | Rv0767c | Rv0767c | 13.389893 |
| 123 | Rv1344 | mbtL | 13.37666 |
| 124 | Rv0848 | cysK2 | 13.308091 |
| 125 | Rv0619 | galTb | 13.238272 |
| 126 | Rv2341 | lppQ | 13.2290945 |
| 127 | Rv0160c | PE4 | 13.080044 |
| 128 | Rv2804c | Rv2804c | 12.992268 |
| 129 | Rv1035c | Rv1035c | 12.986751 |
| 130 | Rv1220c | Rv1220c | 12.913254 |
| 131 | Rv1550 | fadD11 | 12.899639 |
| 132 | Rv1499 | Rv1499 | 12.839415 |
| 133 | Rv0375c | Rv0375c | 12.7841625 |
| 134 | Rv1516c | Rv1516c | 12.755384 |
| 135 | Rv2737A | Rv2737A | 12.743037 |
| 136 | Rv3511 | PE_PGRS55 | 12.729039 |
| 137 | Rv3192 | Rv3192 | 12.704461 |
| 138 | Rv2422 | Rv2422 | 12.692648 |
| 139 | Rv3657c | Rv3657c | 12.687474 |
| 140 | Rv1967 | mce3B | 12.68107 |
| 141 | Rv0620 | galK | 12.59597 |
| 142 | Rv2252 | Rv2252 | 12.577283 |
| 143 | Rv3447c | eccC4 | 12.572819 |
| 144 | Rv1935c | echA13 | 12.510695 |
| 145 | Rv0888 | Rv0888 | 12.428582 |
| 146 | Rv3083 | Rv3083 | 12.409438 |
| 147 | Rv0629c | recD | 12.366861 |
| 148 | Rv1088 | PE9 | 12.3088455 |
| 149 | Rv2352c | PPE38 | 12.2572155 |
| 150 | Rv3506 | fadD17 | 12.2291565 |
| 151 | Rv1972 | Rv1972 | 12.161914 |
| 152 | Rv1706c | PPE23 | 12.108827 |
| 153 | Rv3122 | Rv3122 | 12.074257 |
| 154 | Rv3018A | PE27A | 12.072342 |
| 155 | Rv3468c | Rv3468c | 12.0464735 |
| 156 | Rv2254c | Rv2254c | 12.01951 |
| 157 | Rv3008 | Rv3008 | 11.925177 |
| 158 | Rv0093c | Rv0093c | 11.902434 |
| 159 | Rv1526c | Rv1526c | 11.880957 |
| 160 | Rv2274c | mazF8 | 11.867324 |
| 161 | Rv2843 | Rv2843 | 11.860944 |
| 162 | Rv1946c | lppG | 11.842764 |
| 163 | Rv2351c | plcA | 11.804289 |
| 164 | Rv1355c | moeY | 11.774165 |
| 165 | Rv2250c | Rv2250c | 11.709721 |
| 166 | Rv3181c | Rv3181c | 11.498991 |
| 167 | Rv1482c | Rv1482c | 11.462027 |
| 168 | Rv2897c | Rv2897c | 11.434044 |
| 169 | Rv2997 | Rv2997 | 11.430014 |
| 170 | Rv0912 | Rv0912 | 11.366966 |
| 171 | Rv2654c | Rv2654c | 11.294941 |
| 172 | Rv2769c | PE27 | 11.270819 |
| 173 | Rv1921c | lppF | 11.207877 |
| 174 | Rv3353c | Rv3353c | 11.203488 |
| 175 | Rv3262 | fbiB | 11.195241 |
| 176 | Rv3585 | radA | 11.124298 |
| 177 | Rv3911 | sigM | 11.121451 |
| 178 | Rv0851c | Rv0851c | 11.062332 |
| 179 | Rv1767 | Rv1767 | 10.974101 |
| 180 | Rv0662c | vapB7 | 10.916268 |
| 181 | Rv2506 | Rv2506 | 10.915194 |
| 182 | Rv3191c | Rv3191c | 10.864415 |
| 183 | Rv2988c | leuC | 10.771466 |
| 184 | Rv3330 | dacB1 | 10.74984 |
| 185 | Rv1214c | PE14 | 10.696056 |
| 186 | Rv0456B | mazE1 | 10.695671 |
| 187 | Rv2381c | mbtD | 10.675677 |
| 188 | Rv1958c | Rv1958c | 10.625601 |
| 189 | Rv0843 | Rv0843 | 10.605275 |
| 190 | Rv1879 | Rv1879 | 10.581205 |
| 191 | Rv0090 | Rv0090 | 10.504862 |
| 192 | Rv0078 | Rv0078 | 10.490851 |
| 193 | Rv0024 | Rv0024 | 10.386773 |
| 194 | Rv0791c | Rv0791c | 10.37059 |
| 195 | Rv0969 | ctpV | 10.339356 |
| 196 | Rv3512 | PE_PGRS56 | 10.310704 |
| 197 | Rv0109 | PE_PGRS1 | 10.302506 |
| 198 | Rv2893 | Rv2893 | 10.3017435 |
| 199 | Rv1067c | PE_PGRS19 | 10.294628 |
| 200 | Rv3554 | fdxB | 10.28746 |
| 201 | Rv3623 | lpqG | 10.286108 |
| 202 | Rv0596c | vapB4 | 10.264935 |
| 203 | Rv1068c | PE_PGRS20 | 10.204786 |
| 204 | Rv2250A | Rv2250A | 10.169986 |
| 205 | Rv2017 | Rv2017 | 10.152202 |
| 206 | Rv2546 | vapC18 | 10.138137 |
| 207 | Rv3658c | Rv3658c | 9.998689 |
| 208 | Rv2408 | PE24 | 9.969775 |
| 209 | Rv1383 | carA | 9.952406 |
| 210 | Rv2434c | Rv2434c | 9.909851 |
| 211 | Rv0731c | Rv0731c | 9.840221 |
| 212 | Rv2787 | Rv2787 | 9.834816 |
| 213 | Rv2261c | Rv2261c | 9.834276 |
| 214 | Rv3374 | echA18.1 | 9.823434 |
| 215 | Rv2407 | Rv2407 | 9.813153 |
| 216 | Rv2917 | Rv2917 | 9.790218 |
| 217 | Rv0196 | Rv0196 | 9.773797 |
| 218 | Rv1347c | mbtK | 9.769765 |
| 219 | Rv3395c | Rv3395c | 9.762144 |
| 220 | Rv1186c | Rv1186c | 9.760917 |
| 221 | Rv0763c | Rv0763c | 9.752184 |
| 222 | Rv1734c | Rv1734c | 9.74747 |
| 223 | Rv1961 | Rv1961 | 9.742717 |
| 224 | Rv0536 | galE3 | 9.739242 |
| 225 | Rv0725c | Rv0725c | 9.729004 |
| 226 | Rv1050 | Rv1050 | 9.724152 |
| 227 | Rv2236c | cobD | 9.702332 |
| 228 | Rv1561 | vapC11 | 9.682345 |
| 229 | Rv0448c | Rv0448c | 9.634894 |
| 230 | Rv0219 | Rv0219 | 9.593696 |
| 231 | Rv1395 | Rv1395 | 9.589032 |
| 232 | Rv1469 | ctpD | 9.51175 |
| 233 | Rv2871 | vapB43 | 9.4797735 |
| 234 | Rv1735c | Rv1735c | 9.470944 |
| 235 | Rv3667 | acs | 9.44027 |
| 236 | Rv3055 | Rv3055 | 9.429994 |
| 237 | Rv3373 | echA18 | 9.41571 |
| 238 | Rv0144 | Rv0144 | 9.414421 |
| 239 | Rv0841 | Rv0841 | 9.391798 |
| 240 | Rv1774 | Rv1774 | 9.376637 |
| 241 | Rv0904c | accD3 | 9.333484 |
| 242 | Rv1048c | Rv1048c | 9.278179 |
| 243 | Rv0849 | Rv0849 | 9.266259 |
| 244 | Rv3664c | dppC | 9.246043 |
| 245 | Rv1934c | fadE17 | 9.245542 |
| 246 | Rv3743c | ctpJ | 9.231452 |
| 247 | Rv0259c | Rv0259c | 9.225657 |
| 248 | Rv2812 | Rv2812 | 9.224766 |
| 249 | Rv1853 | ureD | 9.201286 |
| 250 | Rv1663 | pks17 | 9.154806 |
| 251 | Rv1858 | modB | 9.145952 |
| 252 | Rv0550c | vapB3 | 9.134158 |
| 253 | Rv0306 | Rv0306 | 9.108091 |
| 254 | Rv3012c | gatC | 9.106117 |
| 255 | Rv0812 | Rv0812 | 9.081064 |
| 256 | Rv3745c | Rv3745c | 9.075097 |
| 257 | Rv2573 | Rv2573 | 9.065614 |
| 258 | Rv2600 | Rv2600 | 9.050851 |
| 259 | Rv3714c | Rv3714c | 9.042637 |
| 260 | Rv0793 | Rv0793 | 9.019358 |
| 261 | Rv3622c | PE32 | 8.994924 |
| 262 | Rv2300c | Rv2300c | 8.981389 |
| 263 | Rv0971c | echA7 | 8.867619 |
| 264 | Rv3640c | Rv3640c | 8.841994 |
| 265 | Rv0209 | Rv0209 | 8.837655 |
| 266 | Rv3189 | Rv3189 | 8.834909 |
| 267 | Rv3756c | proZ | 8.808011 |
| 268 | Rv0976c | Rv0976c | 8.727799 |
| 269 | Rv0378 | Rv0378 | 8.707435 |
| 270 | Rv0943c | Rv0943c | 8.703175 |
| 271 | Rv3379c | dxs2 | 8.68353 |
| 272 | Rv0122 | Rv0122 | 8.677701 |
| 273 | Rv0792c | Rv0792c | 8.640308 |
| 274 | Rv0197 | Rv0197 | 8.637885 |
| 275 | Rv1456c | Rv1456c | 8.631548 |
| 276 | Rv1225c | Rv1225c | 8.630263 |
| 277 | Rv1973 | Rv1973 | 8.629382 |
| 278 | Rv0063 | Rv0063 | 8.620082 |
| 279 | Rv1403c | Rv1403c | 8.598649 |
| 280 | Rv3204 | Rv3204 | 8.583106 |
| 281 | Rv0403c | mmpS1 | 8.552631 |
| 282 | Rv2864c | Rv2864c | 8.55129 |
| 283 | Rv3135 | PPE50 | 8.534555 |
| 284 | Rv1721c | vapB12 | 8.501909 |
| 285 | Rv3809c | glf | 8.468828 |
| 286 | Rv1005c | pabB | 8.464531 |
| 287 | Rv1227c | Rv1227c | 8.430689 |
| 288 | Rv2119 | Rv2119 | 8.363993 |
| 289 | Rv2415c | Rv2415c | 8.356378 |
| 290 | Rv0650 | Rv0650 | 8.349637 |
| 291 | Rv0105c | rpmB1 | 8.347967 |
| 292 | Rv0034 | Rv0034 | 8.325617 |
| 293 | Rv0751c | mmsB | 8.296175 |
| 294 | Rv0840c | pip | 8.283919 |
| 295 | Rv0310c | Rv0310c | 8.220696 |
| 296 | Rv1116 | Rv1116 | 8.21322 |
| 297 | Rv1552 | frdA | 8.145045 |
| 298 | Rv1519 | Rv1519 | 8.130028 |
| 299 | Rv3366 | spoU | 8.118893 |
| 300 | Rv1999c | Rv1999c | 8.116476 |
| 301 | Rv0494 | Rv0494 | 8.113611 |
| 302 | Rv2180c | Rv2180c | 8.112387 |
| 303 | Rv0274 | Rv0274 | 8.094635 |
| 304 | Rv2747 | argA | 8.044785 |
| 305 | Rv1968 | mce3C | 8.023539 |
| 306 | Rv1372 | Rv1372 | 8.0099325 |
| 307 | Rv1864c | Rv1864c | 8.006814 |
| 308 | Rv2672 | Rv2672 | 7.975011 |
| 309 | Rv0790c | Rv0790c | 7.965493 |
| 310 | Rv2333c | stp | 7.9461727 |
| 311 | Rv2485c | lipQ | 7.9461517 |
| 312 | Rv0119 | fadD7 | 7.90628 |
| 313 | Rv3394c | Rv3394c | 7.894706 |
| 314 | Rv2813 | Rv2813 | 7.872486 |
| 315 | Rv0963c | Rv0963c | 7.8713346 |
| 316 | Rv3344c | PE_PGRS49 | 7.8661814 |
| 317 | Rv1385 | pyrF | 7.861874 |
| 318 | Rv1537 | dinX | 7.860974 |
| 319 | Rv0224c | Rv0224c | 7.8536453 |
| 320 | Rv1651c | PE_PGRS30 | 7.8474593 |
| 321 | Rv2615c | PE_PGRS45 | 7.8446107 |
| 322 | Rv3013 | Rv3013 | 7.8362846 |
| 323 | Rv3448 | eccD4 | 7.8294277 |
| 324 | Rv3510c | Rv3510c | 7.8189144 |
| 325 | Rv0447c | ufaA1 | 7.774213 |
| 326 | Rv0549c | vapC3 | 7.768712 |
| 327 | Rv1714 | Rv1714 | 7.741935 |
| 328 | Rv3555c | Rv3555c | 7.732873 |
| 329 | Rv3419c | gcp | 7.7294707 |
| 330 | Rv0962c | lprP | 7.7016344 |
| 331 | Rv0115 | hddA | 7.665542 |
| 332 | Rv0077c | Rv0077c | 7.649855 |
| 333 | Rv0601c | Rv0601c | 7.6357117 |
| 334 | Rv3178 | Rv3178 | 7.630155 |
| 335 | Rv1760 | Rv1760 | 7.5865817 |
| 336 | Rv3855 | ethR | 7.57502 |
| 337 | Rv3563 | fadE32 | 7.5607147 |
| 338 | Rv2275 | Rv2275 | 7.550774 |
| 339 | Rv1120c | Rv1120c | 7.550562 |
| 340 | Rv2324 | Rv2324 | 7.547983 |
| 341 | Rv3074 | Rv3074 | 7.543722 |
| 342 | Rv1204c | Rv1204c | 7.540247 |
| 343 | Rv3695 | Rv3695 | 7.5276036 |
| 344 | Rv1702c | Rv1702c | 7.5211864 |
| 345 | Rv3384c | vapC46 | 7.5178347 |
| 346 | Rv2011c | Rv2011c | 7.500847 |
| 347 | Rv1041c | Rv1041c | 7.4642215 |
| 348 | Rv0891c | Rv0891c | 7.444831 |
| 349 | Rv3444c | esxT | 7.436032 |
| 350 | Rv2647 | Rv2647 | 7.4202023 |
| 351 | Rv3076 | Rv3076 | 7.4081407 |
| 352 | Rv0699 | Rv0699 | 7.3909073 |
| 353 | Rv2767c | Rv2767c | 7.384072 |
| 354 | Rv2025c | Rv2025c | 7.374321 |
| 355 | Rv1008 | tatD | 7.349339 |
| 356 | Rv2158c | murE | 7.311332 |
| 357 | Rv2763c | dfrA | 7.292768 |
| 358 | Rv1592c | Rv1592c | 7.2585597 |
| 359 | Rv3370c | dnaE2 | 7.248392 |
| 360 | Rv2099c | PE21 | 7.2215652 |
| 361 | Rv3731 | ligC | 7.202317 |
| 362 | Rv1676 | Rv1676 | 7.1966004 |
| 363 | Rv0630c | recB | 7.177355 |
| 364 | Rv2469c | Rv2469c | 7.160962 |
| 365 | Rv0322 | udgA | 7.1571016 |
| 366 | Rv2000 | Rv2000 | 7.1452045 |
| 367 | Rv1811 | mgtC | 7.123348 |
| 368 | Rv0414c | thiE | 7.1155868 |
| 369 | Rv0613c | Rv0613c | 7.1099653 |
| 370 | Rv3660c | Rv3660c | 7.1069345 |
| 371 | Rv2489c | Rv2489c | 7.1039276 |
| 372 | Rv3573c | fadE34 | 7.0873528 |
| 373 | Rv3201c | Rv3201c | 7.071653 |
| 374 | Rv2661c | Rv2661c | 7.0316987 |
| 375 | Rv1150 | Rv1150 | 7.011491 |
| 376 | Rv0394c | Rv0394c | 7.00648 |
| 377 | Rv3470c | ilvB2 | 7.0056057 |
| 378 | Rv2235 | Rv2235 | 7.0045547 |
| 379 | Rv2944 | Rv2944 | 6.997806 |
| 380 | Rv0324 | Rv0324 | 6.9820614 |
| 381 | Rv2331 | Rv2331 | 6.9770966 |
| 382 | Rv2230c | Rv2230c | 6.9717474 |
| 383 | Rv2779c | Rv2779c | 6.9671946 |
| 384 | Rv2655c | Rv2655c | 6.964982 |
| 385 | Rv2436 | rbsK | 6.95012 |
| 386 | Rv3428c | Rv3428c | 6.9392686 |
| 387 | Rv2567 | Rv2567 | 6.9275937 |
| 388 | Rv0150c | Rv0150c | 6.9091053 |
| 389 | Rv3337 | Rv3337 | 6.900306 |
| 390 | Rv0162c | adhE1 | 6.8839335 |
| 391 | Rv2251 | Rv2251 | 6.8828073 |
| 392 | Rv1258c | Rv1258c | 6.87072 |
| 393 | Rv2423 | Rv2423 | 6.867015 |
| 394 | Rv3022A | PE29 | 6.866856 |
| 395 | Rv1730c | Rv1730c | 6.8635216 |
| 396 | Rv3649 | Rv3649 | 6.862316 |
| 397 | Rv3333c | Rv3333c | 6.844626 |
| 398 | Rv2001 | Rv2001 | 6.83854 |
| 399 | Rv2862A | vapB23 | 6.7963777 |
| 400 | Rv0067c | Rv0067c | 6.7876124 |
| 401 | Rv3307 | deoD | 6.767349 |
| 402 | Rv3402c | Rv3402c | 6.7571063 |
| 403 | Rv2656c | Rv2656c | 6.755376 |
| 404 | Rv0319 | pcp | 6.751636 |
| 405 | Rv1674c | Rv1674c | 6.741805 |
| 406 | Rv3445c | esxU | 6.7413807 |
| 407 | Rv3397c | phyA | 6.7395997 |
| 408 | Rv2077A | Rv2077A | 6.7284846 |
| 409 | Rv1966 | mce3A | 6.7140193 |
| 410 | Rv2188c | pimB | 6.7026157 |
| 411 | Rv1430 | PE16 | 6.701857 |
| 412 | Rv3905c | esxF | 6.6712275 |
| 413 | Rv1087A | Rv1087A | 6.6676598 |
| 414 | Rv3741c | Rv3741c | 6.665955 |
| 415 | Rv1045 | Rv1045 | 6.6645694 |
| 416 | Rv0737 | Rv0737 | 6.6594467 |
| 417 | Rv1928c | Rv1928c | 6.6454034 |
| 418 | Rv1381 | pyrC | 6.6399136 |
| 419 | Rv2758c | vapB21 | 6.638484 |
| 420 | Rv0260c | Rv0260c | 6.6228023 |
| 421 | Rv0139 | Rv0139 | 6.6174054 |
| 422 | Rv3093c | Rv3093c | 6.6168356 |
| 423 | Rv1433 | Rv1433 | 6.607271 |
| 424 | Rv3422c | Rv3422c | 6.602184 |
| 425 | Rv2293c | Rv2293c | 6.5775933 |
| 426 | Rv2082 | Rv2082 | 6.5754924 |
| 427 | Rv2892c | PPE45 | 6.548107 |
| 428 | Rv1189 | sigI | 6.5312085 |
| 429 | Rv0330c | Rv0330c | 6.517551 |
| 430 | Rv0358 | Rv0358 | 6.484505 |
| 431 | Rv2107 | PE22 | 6.4792743 |
| 432 | Rv3654c | Rv3654c | 6.457265 |
| 433 | Rv0576 | Rv0576 | 6.4498878 |
| 434 | Rv0964c | Rv0964c | 6.4362535 |
| 435 | Rv2427A | oxyR' | 6.421873 |
| 436 | Rv1252c | lprE | 6.4165573 |
| 437 | Rv1965 | yrbE3B | 6.411837 |
| 438 | Rv2282c | Rv2282c | 6.405134 |
| 439 | Rv0226c | Rv0226c | 6.4045067 |
| 440 | Rv3906c | Rv3906c | 6.4040046 |
| 441 | Rv0974c | accD2 | 6.370946 |
| 442 | Rv2063A | mazF7 | 6.368206 |
| 443 | Rv3557c | Rv3557c | 6.358317 |
| 444 | Rv3812 | PE_PGRS62 | 6.3542504 |
| 445 | Rv3538 | Rv3538 | 6.3492384 |
| 446 | Rv2596 | vapC40 | 6.3481736 |
| 447 | Rv1349 | irtB | 6.3475127 |
| 448 | Rv2012 | Rv2012 | 6.3280807 |
| 449 | Rv2309A | Rv2309A | 6.327676 |
| 450 | Rv2645 | Rv2645 | 6.3223085 |
| 451 | Rv0612 | Rv0612 | 6.3166146 |
| 452 | Rv3770A | Rv3770A | 6.315118 |
| 453 | Rv2292c | Rv2292c | 6.307998 |
| 454 | Rv0745 | Rv0745 | 6.3035035 |
| 455 | Rv3065 | mmr | 6.300322 |
| 456 | Rv3425 | PPE57 | 6.2772717 |
| 457 | Rv1706A | Rv1706A | 6.2456255 |
| 458 | Rv0735 | sigL | 6.241223 |
| 459 | Rv3026c | Rv3026c | 6.2402487 |
| 460 | Rv2268c | cyp128 | 6.213489 |
| 461 | Rv1725c | Rv1725c | 6.210119 |
| 462 | Rv3351c | Rv3351c | 6.200558 |
| 463 | Rv2392 | cysH | 6.187223 |
| 464 | Rv0602c | tcrA | 6.1770706 |
| 465 | Rv3226c | Rv3226c | 6.1770644 |
| 466 | Rv0647c | Rv0647c | 6.1760154 |
| 467 | Rv3469c | mhpE | 6.1704755 |
| 468 | Rv1111c | Rv1111c | 6.170438 |
| 469 | Rv2225 | panB | 6.1555986 |
| 470 | Rv3822 | Rv3822 | 6.1535277 |
| 471 | Rv0071 | Rv0071 | 6.143785 |
| 472 | Rv2160c | Rv2160c | 6.1299043 |
| 473 | Rv3521 | Rv3521 | 6.1242905 |
| 474 | Rv3564 | fadE33 | 6.1107507 |
| 475 | Rv0980c | PE_PGRS18 | 6.1011996 |
| 476 | Rv0915c | PPE14 | 6.090141 |
| 477 | Rv3817 | Rv3817 | 6.077533 |
| 478 | Rv3565 | aspB | 6.0769334 |
| 479 | Rv2089c | pepE | 6.0659156 |
| 480 | Rv1103c | mazE3 | 6.064738 |
| 481 | Rv1673c | Rv1673c | 6.0385513 |
| 482 | Rv0817c | Rv0817c | 6.0226874 |
| 483 | Rv2232 | ptkA | 6.0027485 |
| 484 | Rv3904c | esxE | 6.001965 |
| 485 | Rv0493c | Rv0493c | 6.001695 |
| 486 | Rv3426 | PPE58 | 5.996001 |
| 487 | Rv0959A | vapB9 | 5.984733 |
| 488 | Rv3129 | Rv3129 | 5.9817944 |
| 489 | Rv0753c | mmsA | 5.976074 |
| 490 | Rv3532 | PPE61 | 5.972012 |
| 491 | Rv1807 | PPE31 | 5.9703875 |
| 492 | Rv1492 | mutA | 5.9496703 |
| 493 | Rv2308 | Rv2308 | 5.909527 |
| 494 | Rv0210 | Rv0210 | 5.9056745 |
| 495 | Rv2063 | mazE7 | 5.8981447 |
| 496 | Rv1226c | Rv1226c | 5.889888 |
| 497 | Rv3718c | Rv3718c | 5.8886404 |
| 498 | Rv2438A | Rv2438A | 5.8859425 |
| 499 | Rv0258c | Rv0258c | 5.8836083 |
| 500 | Rv0836c | Rv0836c | 5.879046 |
| 501 | Rv1426c | lipO | 5.877169 |
| 502 | Rv0527 | ccdA | 5.869602 |
| 503 | Rv0607 | Rv0607 | 5.8542304 |
| 504 | Rv2589 | gabT | 5.8512 |
| 505 | Rv0752c | fadE9 | 5.843796 |
| 506 | Rv1873 | Rv1873 | 5.841243 |
| 507 | Rv1551 | plsB1 | 5.830927 |
| 508 | Rv1136 | Rv1136 | 5.827624 |
| 509 | Rv1597 | Rv1597 | 5.816356 |
| 510 | Rv0444c | rskA | 5.7936506 |
| 511 | Rv3391 | acrA1 | 5.7902746 |
| 512 | Rv0787 | Rv0787 | 5.778405 |
| 513 | Rv0754 | PE_PGRS11 | 5.766983 |
| 514 | Rv1927 | Rv1927 | 5.760133 |
| 515 | Rv2123 | PPE37 | 5.755066 |
| 516 | Rv1244 | lpqZ | 5.747441 |
| 517 | Rv2618 | Rv2618 | 5.729051 |
| 518 | Rv0560c | Rv0560c | 5.7280674 |
| 519 | Rv1866 | Rv1866 | 5.727995 |
| 520 | Rv0388c | PPE9 | 5.7270985 |
| 521 | Rv2759c | vapC42 | 5.722149 |
| 522 | Rv0779c | Rv0779c | 5.7219086 |
| 523 | Rv0381c | Rv0381c | 5.7033143 |
| 524 | Rv1717 | Rv1717 | 5.6959953 |
| 525 | Rv0179c | lprO | 5.6878366 |
| 526 | Rv1748 | Rv1748 | 5.6826606 |
| 527 | Rv0561c | Rv0561c | 5.676792 |
| 528 | Rv2768c | PPE43 | 5.665625 |
| 529 | Rv1434 | Rv1434 | 5.66218 |
| 530 | Rv0163 | Rv0163 | 5.6599674 |
| 531 | Rv0492c | Rv0492c | 5.630256 |
| 532 | Rv3034c | Rv3034c | 5.62341 |
| 533 | Rv0775 | Rv0775 | 5.6231146 |
| 534 | Rv2913c | Rv2913c | 5.6218 |
| 535 | Rv1685c | Rv1685c | 5.602895 |
| 536 | Rv0691c | Rv0691c | 5.5894556 |
| 537 | Rv1354c | Rv1354c | 5.582676 |
| 538 | Rv0956 | purN | 5.5803847 |
| 539 | Rv0252 | nirB | 5.577752 |
| 540 | Rv1684 | Rv1684 | 5.5716224 |
| 541 | Rv3454 | Rv3454 | 5.5676117 |
| 542 | Rv1781c | malQ | 5.5654306 |
| 543 | Rv3259 | Rv3259 | 5.5643353 |
| 544 | Rv0104 | Rv0104 | 5.5617 |
| 545 | Rv2320c | rocE | 5.5535183 |
| 546 | Rv3909 | Rv3909 | 5.5521584 |
| 547 | Rv1727 | Rv1727 | 5.5521374 |
| 548 | Rv1272c | Rv1272c | 5.5422244 |
| 549 | Rv3073c | Rv3073c | 5.5421567 |
| 550 | Rv2730 | Rv2730 | 5.5344486 |
| 551 | Rv1615 | Rv1615 | 5.533779 |
| 552 | Rv2179c | Rv2179c | 5.5314384 |
| 553 | Rv2607 | pdxH | 5.514571 |
| 554 | Rv0371c | Rv0371c | 5.5143533 |
| 555 | Rv1414 | Rv1414 | 5.50941 |
| 556 | Rv2331A | Rv2331A | 5.50753 |
| 557 | Rv3415c | Rv3415c | 5.5060186 |
| 558 | Rv3851 | Rv3851 | 5.4989586 |
| 559 | Rv1897c | Rv1897c | 5.4972706 |
| 560 | Rv0535 | pnp | 5.495184 |
| 561 | Rv0789c | Rv0789c | 5.486773 |
| 562 | Rv1051c | Rv1051c | 5.47601 |
| 563 | Rv1295 | thrC | 5.475008 |
| 564 | Rv2231c | cobC | 5.47474 |
| 565 | Rv1913 | Rv1913 | 5.453473 |
| 566 | Rv0676c | mmpL5 | 5.4469886 |
| 567 | Rv2508c | Rv2508c | 5.445852 |
| 568 | Rv2859c | Rv2859c | 5.445689 |
| 569 | Rv3170 | aofH | 5.4394917 |
| 570 | Rv2227 | Rv2227 | 5.438764 |
| 571 | Rv0205 | Rv0205 | 5.4370213 |
| 572 | Rv2691 | ceoB | 5.433415 |
| 573 | Rv2547 | vapB19 | 5.4317656 |
| 574 | Rv1200 | Rv1200 | 5.428075 |
| 575 | Rv2670c | Rv2670c | 5.423671 |
| 576 | Rv3382c | lytB1 | 5.4227905 |
| 577 | Rv0526 | Rv0526 | 5.410534 |
| 578 | Rv0508 | Rv0508 | 5.395212 |
| 579 | Rv0854 | Rv0854 | 5.39318 |
| 580 | Rv1327c | glgE | 5.3835936 |
| 581 | Rv0776c | Rv0776c | 5.3581867 |
| 582 | Rv1634 | Rv1634 | 5.3568683 |
| 583 | Rv0551c | fadD8 | 5.35673 |
| 584 | Rv3643 | Rv3643 | 5.3533063 |
| 585 | Rv1112 | Rv1112 | 5.347266 |
| 586 | Rv0161 | Rv0161 | 5.339634 |
| 587 | Rv2281 | pitB | 5.3339667 |
| 588 | Rv0225 | Rv0225 | 5.32819 |
| 589 | Rv2835c | ugpA | 5.3237247 |
| 590 | Rv0255c | cobQ1 | 5.3178477 |
| 591 | Rv0916c | PE7 | 5.313343 |
| 592 | Rv1075c | Rv1075c | 5.304822 |
| 593 | Rv0267 | narU | 5.3026323 |
| 594 | Rv0588 | yrbE2B | 5.2877307 |
| 595 | Rv1010 | ksgA | 5.28609 |
| 596 | Rv3439c | Rv3439c | 5.267671 |
| 597 | Rv0594 | mce2F | 5.2629805 |
| 598 | Rv1291c | Rv1291c | 5.2621984 |
| 599 | Rv0158 | Rv0158 | 5.258882 |
| 600 | Rv0837c | Rv0837c | 5.251964 |
| 601 | Rv1455 | Rv1455 | 5.2473836 |
| 602 | Rv1546 | Rv1546 | 5.244122 |
| 603 | Rv3740c | Rv3740c | 5.2366867 |
| 604 | Rv0830 | Rv0830 | 5.2271495 |
| 605 | Rv1157c | Rv1157c | 5.217875 |
| 606 | Rv3349c | Rv3349c | 5.20682 |
| 607 | Rv0521 | Rv0521 | 5.2058764 |
| 608 | Rv1353c | Rv1353c | 5.195502 |
| 609 | Rv3504 | fadE26 | 5.185068 |
| 610 | Rv2722 | Rv2722 | 5.1810117 |
| 611 | Rv0428c | Rv0428c | 5.1757536 |
| 612 | Rv1203c | Rv1203c | 5.164 |
| 613 | Rv1158c | Rv1158c | 5.1626124 |
| 614 | Rv2848c | cobB | 5.1540804 |
| 615 | Rv3182 | Rv3182 | 5.148729 |
| 616 | Rv3797 | fadE35 | 5.1470037 |
| 617 | Rv2961 | Rv2961 | 5.133844 |
| 618 | Rv2566 | Rv2566 | 5.1304398 |
| 619 | Rv3085 | Rv3085 | 5.121445 |
| 620 | Rv0130 | htdZ | 5.108442 |
| 621 | Rv2395 | Rv2395 | 5.1016183 |
| 622 | Rv2092c | helY | 5.099019 |
| 623 | Rv2456c | Rv2456c | 5.091326 |
| 624 | Rv2642 | Rv2642 | 5.085983 |
| 625 | Rv2316 | uspA | 5.083272 |
| 626 | Rv3398c | idsA1 | 5.072272 |
| 627 | Rv1319c | Rv1319c | 5.0647407 |
| 628 | Rv3793 | embC | 5.0567923 |
| 629 | Rv1119c | Rv1119c | 5.054762 |
| 630 | Rv0519c | Rv0519c | 5.053383 |
| 631 | Rv2022c | Rv2022c | 5.002853 |
| 632 | Rv2046 | lppI | 4.9945326 |
| 633 | Rv0882 | Rv0882 | 4.982498 |
| 634 | Rv3580c | cysS1 | 4.968428 |
| 635 | Rv1485 | hemZ | 4.963308 |
| 636 | Rv2943 | Rv2943 | 4.9588804 |
| 637 | Rv3737 | Rv3737 | 4.948565 |
| 638 | Rv2458 | mmuM | 4.945776 |
| 639 | Rv0185 | Rv0185 | 4.9277897 |
| 640 | Rv0328 | Rv0328 | 4.924178 |
| 641 | Rv2786c | ribF | 4.9172072 |
| 642 | Rv0329c | Rv0329c | 4.9169936 |
| 643 | Rv2287 | yjcE | 4.907734 |
| 644 | Rv3092c | Rv3092c | 4.902658 |
| 645 | Rv2490c | PE_PGRS43 | 4.90182 |
| 646 | Rv3357 | relJ | 4.8995843 |
| 647 | Rv0482 | murB | 4.899147 |
| 648 | Rv2963 | Rv2963 | 4.892181 |
| 649 | Rv1819c | bacA | 4.8915124 |
| 650 | Rv1936 | Rv1936 | 4.882717 |
| 651 | Rv2072c | cobL | 4.8778944 |
| 652 | Rv3160c | Rv3160c | 4.874823 |
| 653 | Rv2757c | vapC21 | 4.8601546 |
| 654 | Rv1242 | vapC33 | 4.8550477 |
| 655 | Rv2209 | Rv2209 | 4.848691 |
| 656 | Rv2801A | mazE9 | 4.8478646 |
| 657 | Rv1235 | lpqY | 4.834536 |
| 658 | Rv1910c | Rv1910c | 4.833365 |
| 659 | Rv3757c | proW | 4.833085 |
| 660 | Rv2472 | Rv2472 | 4.8281384 |
| 661 | Rv1716 | Rv1716 | 4.824783 |
| 662 | Rv0960 | vapC9 | 4.819355 |
| 663 | Rv2379c | mbtF | 4.8159266 |
| 664 | Rv1570 | bioD | 4.8142843 |
| 665 | Rv2491 | Rv2491 | 4.8126817 |
| 666 | Rv0215c | fadE3 | 4.810514 |
| 667 | Rv2085 | Rv2085 | 4.8050466 |
| 668 | Rv1473A | Rv1473A | 4.8034487 |
| 669 | Rv1669 | Rv1669 | 4.8021913 |
| 670 | Rv0228 | Rv0228 | 4.7899876 |
| 671 | Rv1003 | Rv1003 | 4.789975 |
| 672 | Rv0552 | Rv0552 | 4.786364 |
| 673 | Rv3860 | Rv3860 | 4.783017 |
| 674 | Rv1325c | PE_PGRS24 | 4.753194 |
| 675 | Rv0117 | oxyS | 4.7483516 |
| 676 | Rv2640c | Rv2640c | 4.7447267 |
| 677 | Rv1523 | Rv1523 | 4.741919 |
| 678 | Rv2113 | Rv2113 | 4.7364826 |
| 679 | Rv1931c | Rv1931c | 4.7290187 |
| 680 | Rv1212c | glgA | 4.72652 |
| 681 | Rv1715 | fadB3 | 4.7262483 |
| 682 | Rv2157c | murF | 4.725993 |
| 683 | Rv3090 | Rv3090 | 4.72585 |
| 684 | Rv2989 | Rv2989 | 4.7192326 |
| 685 | Rv2294 | Rv2294 | 4.716115 |
| 686 | Rv0539 | Rv0539 | 4.710961 |
| 687 | Rv3387 | Rv3387 | 4.707042 |
| 688 | Rv2432c | Rv2432c | 4.7038107 |
| 689 | Rv1867 | Rv1867 | 4.7027144 |
| 690 | Rv3849 | espR | 4.7019353 |
| 691 | Rv0825c | Rv0825c | 4.699813 |
| 692 | Rv1043c | Rv1043c | 4.698978 |
| 693 | Rv0616c | Rv0616c | 4.6964846 |
| 694 | Rv1952 | vapB14 | 4.691324 |
| 695 | Rv2116 | lppK | 4.6887436 |
| 696 | Rv1969 | mce3D | 4.679036 |
| 697 | Rv0095c | Rv0095c | 4.6689334 |
| 698 | Rv0377 | Rv0377 | 4.668662 |
| 699 | Rv0554 | bpoC | 4.66116 |
| 700 | Rv3294c | Rv3294c | 4.6609297 |
| 701 | Rv3304 | Rv3304 | 4.65608 |
| 702 | Rv3032 | Rv3032 | 4.6559935 |
| 703 | Rv3767c | Rv3767c | 4.6475263 |
| 704 | Rv3031 | Rv3031 | 4.6443224 |
| 705 | Rv2317 | uspB | 4.6358976 |
| 706 | Rv1895 | Rv1895 | 4.6251154 |
| 707 | Rv3015c | Rv3015c | 4.6221256 |
| 708 | Rv2595 | vapB40 | 4.6130743 |
| 709 | Rv3126c | Rv3126c | 4.612501 |
| 710 | Rv1049 | Rv1049 | 4.6037645 |
| 711 | Rv3713 | cobQ2 | 4.6030593 |
| 712 | Rv0924c | mntH | 4.591815 |
| 713 | Rv0265c | Rv0265c | 4.591758 |
| 714 | Rv3542c | Rv3542c | 4.588704 |
| 715 | Rv3054c | Rv3054c | 4.5786533 |
| 716 | Rv1062 | Rv1062 | 4.5748415 |
| 717 | Rv2486 | echA14 | 4.5744166 |
| 718 | Rv2719c | Rv2719c | 4.5716295 |
| 719 | Rv2548 | vapC19 | 4.5655437 |
| 720 | Rv1632c | Rv1632c | 4.561008 |
| 721 | Rv2577 | Rv2577 | 4.5589743 |
| 722 | Rv2964 | purU | 4.557643 |
| 723 | Rv3500c | yrbE4B | 4.556882 |
| 724 | Rv1768 | PE_PGRS31 | 4.556721 |
| 725 | Rv1233c | Rv1233c | 4.5497274 |
| 726 | Rv1562c | treZ | 4.5471053 |
| 727 | Rv2433c | Rv2433c | 4.5432324 |
| 728 | Rv3590c | PE_PGRS58 | 4.542777 |
| 729 | Rv2991 | Rv2991 | 4.5336833 |
| 730 | Rv0369c | Rv0369c | 4.531174 |
| 731 | Rv3340 | metC | 4.5308867 |
| 732 | Rv3066 | Rv3066 | 4.528256 |
| 733 | Rv0031 | Rv0031 | 4.525666 |
| 734 | Rv1805c | Rv1805c | 4.5219812 |
| 735 | Rv3647c | Rv3647c | 4.521935 |
| 736 | Rv2451 | Rv2451 | 4.521033 |
| 737 | Rv2578c | Rv2578c | 4.5148897 |
| 738 | Rv3652 | PE_PGRS60 | 4.513328 |
| 739 | Rv2806 | Rv2806 | 4.5042534 |
| 740 | Rv0449c | Rv0449c | 4.500841 |
| 741 | Rv1348 | irtA | 4.498603 |
| 742 | Rv2161c | Rv2161c | 4.4959927 |
| 743 | Rv0387c | Rv0387c | 4.4879904 |
| 744 | Rv1803c | PE_PGRS32 | 4.487718 |
| 745 | Rv2065 | cobH | 4.487446 |
| 746 | Rv0214 | fadD4 | 4.4837914 |
| 747 | Rv3665c | dppB | 4.480077 |
| 748 | Rv0923c | Rv0923c | 4.479663 |
| 749 | Rv1528c | papA4 | 4.4705133 |
| 750 | Rv0396 | Rv0396 | 4.4635873 |
| 751 | Rv0595c | vapC4 | 4.4615917 |
| 752 | Rv0900 | Rv0900 | 4.4387197 |
| 753 | Rv2253 | Rv2253 | 4.436087 |
| 754 | Rv3507 | PE_PGRS53 | 4.425701 |
| 755 | Rv3070 | Rv3070 | 4.424006 |
| 756 | Rv2854 | Rv2854 | 4.4230337 |
| 757 | Rv2274A | mazE8 | 4.422096 |
| 758 | Rv0216 | Rv0216 | 4.414311 |
| 759 | Rv2998A | Rv2998A | 4.412118 |
| 760 | Rv1963c | mce3R | 4.4096584 |
| 761 | Rv1014c | pth | 4.4000707 |
| 762 | Rv0068 | Rv0068 | 4.399714 |
| 763 | Rv3635 | Rv3635 | 4.399469 |
| 764 | Rv1188 | Rv1188 | 4.397965 |
| 765 | Rv1620c | cydC | 4.3875656 |
| 766 | Rv3857c | Rv3857c | 4.3841844 |
| 767 | Rv2542 | Rv2542 | 4.3771067 |
| 768 | Rv2560 | Rv2560 | 4.374958 |
| 769 | Rv1123c | bpoB | 4.373898 |
| 770 | Rv3347c | PPE55 | 4.361976 |
| 771 | Rv3730c | Rv3730c | 4.360789 |
| 772 | Rv1982c | vapC36 | 4.35832 |
| 773 | Rv3062 | ligB | 4.357623 |
| 774 | Rv2983 | Rv2983 | 4.3550076 |
| 775 | Rv1542c | glbN | 4.3547406 |
| 776 | Rv1058 | fadD14 | 4.3532567 |
| 777 | Rv2828c | Rv2828c | 4.3463273 |
| 778 | Rv2464c | Rv2464c | 4.3446474 |
| 779 | Rv0253 | nirD | 4.3408084 |
| 780 | Rv3840 | Rv3840 | 4.335496 |
| 781 | Rv2449c | Rv2449c | 4.3311877 |
| 782 | Rv1752 | Rv1752 | 4.3300843 |
| 783 | MTB000078 | MTS2823 | 4.330003 |
| 784 | Rv3901c | Rv3901c | 4.3276234 |
| 785 | Rv3102c | ftsE | 4.317649 |
| 786 | MTB000019 | rrs | 4.3164268 |
| 787 | Rv3670 | ephE | 4.3098164 |
| 788 | Rv3194c | Rv3194c | 4.303717 |
| 789 | Rv0294 | tam | 4.2904654 |
| 790 | Rv1393c | Rv1393c | 4.2846293 |
| 791 | Rv2321c | rocD2 | 4.28375 |
| 792 | Rv3854c | ethA | 4.279672 |
| 793 | Rv1612 | trpB | 4.277375 |
| 794 | Rv3098c | Rv3098c | 4.261271 |
| 795 | Rv2811 | Rv2811 | 4.252256 |
| 796 | Rv3751 | Rv3751 | 4.2497783 |
| 797 | Rv1940 | ribA1 | 4.232509 |
| 798 | Rv3061c | fadE22 | 4.2227135 |
| 799 | Rv3305c | amiA1 | 4.218202 |
| 800 | Rv0327c | cyp135A1 | 4.213596 |
| 801 | Rv1263 | amiB2 | 4.213195 |
| 802 | Rv0858c | dapC | 4.2115927 |
| 803 | Rv3836 | Rv3836 | 4.2096815 |
| 804 | Rv3505 | fadE27 | 4.206741 |
| 805 | Rv0604 | lpqO | 4.2050233 |
| 806 | Rv1129c | Rv1129c | 4.2021937 |
| 807 | Rv1834 | lipZ | 4.1924243 |
| 808 | Rv1450c | PE_PGRS27 | 4.187309 |
| 809 | Rv0159c | PE3 | 4.181329 |
| 810 | Rv1267c | embR | 4.179094 |
| 811 | Rv3027c | Rv3027c | 4.1713357 |
| 812 | Rv0518 | Rv0518 | 4.163753 |
| 813 | Rv2327 | Rv2327 | 4.1550817 |
| 814 | Rv2980 | Rv2980 | 4.1543407 |
| 815 | Rv2594c | ruvC | 4.151208 |
| 816 | Rv1441c | PE_PGRS26 | 4.143676 |
| 817 | Rv3039c | echA17 | 4.141389 |
| 818 | Rv0528 | Rv0528 | 4.1348352 |
| 819 | Rv2659c | Rv2659c | 4.1341395 |
| 820 | Rv1549 | fadD11.1 | 4.1320553 |
| 821 | Rv2601A | vapB41 | 4.127319 |
| 822 | Rv1975 | Rv1975 | 4.1265903 |
| 823 | Rv1001 | arcA | 4.109982 |
| 824 | Rv3123 | Rv3123 | 4.1037955 |
| 825 | Rv1888A | Rv1888A | 4.1019263 |
| 826 | Rv3845 | Rv3845 | 4.101714 |
| 827 | Rv3786c | Rv3786c | 4.094388 |
| 828 | Rv2884 | Rv2884 | 4.0942416 |
| 829 | Rv2121c | hisG | 4.083268 |
| 830 | Rv1688 | mpg | 4.0797157 |
| 831 | Rv0921 | Rv0921 | 4.0722175 |
| 832 | Rv0456A | mazF1 | 4.071094 |
| 833 | Rv1568 | bioA | 4.0642076 |
| 834 | Rv2349c | plcC | 4.061907 |
| 835 | Rv1277 | Rv1277 | 4.060531 |
| 836 | Rv0557 | mgtA | 4.056763 |
| 837 | Rv0842 | Rv0842 | 4.046902 |
| 838 | Rv1216c | Rv1216c | 4.043734 |
| 839 | Rv0894 | Rv0894 | 4.042705 |
| 840 | Rv1399c | nlhH | 4.039837 |
| 841 | Rv1668c | Rv1668c | 4.0261273 |
| 842 | Rv1746 | pknF | 4.021385 |
| 843 | Rv2957 | Rv2957 | 4.0198755 |
| 844 | Rv0439c | Rv0439c | 4.018322 |
| 845 | Rv3594 | Rv3594 | 4.0171375 |
| 846 | Rv1366 | Rv1366 | 4.010584 |
| 847 | Rv1954c | Rv1954c | 4.0082006 |
| 848 | Rv2437 | Rv2437 | 4.006971 |
| 849 | Rv1376 | Rv1376 | 4.0027013 |
| 850 | Rv0360c | Rv0360c | 3.9984987 |
| 851 | Rv2078 | Rv2078 | 3.993983 |
| 852 | Rv0344c | lpqJ | 3.9871516 |
| 853 | Rv1407 | fmu | 3.986065 |
| 854 | Rv0862c | Rv0862c | 3.9745026 |
| 855 | Rv1273c | Rv1273c | 3.9723325 |
| 856 | Rv1044 | Rv1044 | 3.970265 |
| 857 | Rv3617 | ephA | 3.9669929 |
| 858 | Rv3399 | Rv3399 | 3.9645898 |
| 859 | Rv2923c | Rv2923c | 3.9629314 |
| 860 | Rv2015c | Rv2015c | 3.9594223 |
| 861 | Rv2922A | acyP | 3.958088 |
| 862 | Rv2827c | Rv2827c | 3.9580877 |
| 863 | Rv3806c | ubiA | 3.955906 |
| 864 | Rv2644c | Rv2644c | 3.95514 |
| 865 | Rv2748c | ftsK | 3.9537091 |
| 866 | Rv3541c | Rv3541c | 3.9529989 |
| 867 | Rv2920c | amt | 3.9461906 |
| 868 | Rv1358 | Rv1358 | 3.9456067 |
| 869 | Rv1591 | Rv1591 | 3.9332047 |
| 870 | Rv1533 | Rv1533 | 3.9329238 |
| 871 | Rv2344c | dgt | 3.929587 |
| 872 | Rv3641c | fic | 3.928335 |
| 873 | Rv3082c | virS | 3.927055 |
| 874 | Rv0187 | Rv0187 | 3.9229138 |
| 875 | Rv1339 | Rv1339 | 3.921717 |
| 876 | Rv2440c | obg | 3.9187891 |
| 877 | Rv3892c | PPE69 | 3.913624 |
| 878 | Rv3721c | dnaZX | 3.9009147 |
| 879 | Rv3106 | fprA | 3.9008763 |
| 880 | Rv0898c | Rv0898c | 3.8999317 |
| 881 | Rv1732c | Rv1732c | 3.8880877 |
| 882 | Rv3608c | folP1 | 3.8876247 |
| 883 | Rv3406 | Rv3406 | 3.8832932 |
| 884 | Rv0165c | mce1R | 3.882199 |
| 885 | Rv2826c | Rv2826c | 3.8803127 |
| 886 | Rv3673c | Rv3673c | 3.8778758 |
| 887 | Rv1625c | cya | 3.877586 |
| 888 | Rv1742 | Rv1742 | 3.8768713 |
| 889 | Rv1089 | PE10 | 3.875954 |
| 890 | Rv3368c | Rv3368c | 3.875863 |
| 891 | Rv1166 | lpqW | 3.875557 |
| 892 | Rv2824c | Rv2824c | 3.8753667 |
| 893 | Rv0481c | Rv0481c | 3.8689418 |
| 894 | Rv3017c | esxQ | 3.8659716 |
| 895 | Rv1218c | Rv1218c | 3.8622115 |
| 896 | Rv1384 | carB | 3.8488595 |
| 897 | Rv0195 | Rv0195 | 3.8486195 |
| 898 | Rv1939 | Rv1939 | 3.8464158 |
| 899 | Rv3744 | nmtR | 3.845553 |
| 900 | Rv3472 | Rv3472 | 3.8450866 |
| 901 | Rv2898c | Rv2898c | 3.8436635 |
| 902 | Rv0152c | PE2 | 3.8409612 |
| 903 | Rv2906c | trmD | 3.840617 |
| 904 | Rv2018 | Rv2018 | 3.837064 |
| 905 | Rv1708 | Rv1708 | 3.8315828 |
| 906 | MTB000016 | glnT | 3.830503 |
| 907 | Rv1090 | celA2b | 3.8251874 |
| 908 | Rv0008c | Rv0008c | 3.819222 |
| 909 | Rv1851 | ureF | 3.816767 |
| 910 | Rv3392c | cmaA1 | 3.8133903 |
| 911 | Rv0756c | Rv0756c | 3.8104873 |
| 912 | Rv1930c | Rv1930c | 3.7954397 |
| 913 | Rv2488c | Rv2488c | 3.7846591 |
| 914 | Rv3655c | Rv3655c | 3.7727625 |
| 915 | Rv2122c | hisE | 3.770518 |
| 916 | Rv2154c | ftsW | 3.7660334 |
| 917 | Rv3095 | Rv3095 | 3.7614157 |
| 918 | Rv1357c | Rv1357c | 3.7579205 |
| 919 | Rv2291 | sseB | 3.7565627 |
| 920 | Rv0261c | narK3 | 3.7489307 |
| 921 | Rv1292 | argS | 3.7395537 |
| 922 | Rv1002c | Rv1002c | 3.7386703 |
| 923 | Rv3485c | Rv3485c | 3.738535 |
| 924 | Rv2515c | Rv2515c | 3.7382712 |
| 925 | Rv1318c | Rv1318c | 3.7374134 |
| 926 | Rv2088 | pknJ | 3.736678 |
| 927 | Rv1947 | Rv1947 | 3.7347002 |
| 928 | Rv0362 | mgtE | 3.7246668 |
| 929 | Rv0945 | Rv0945 | 3.719358 |
| 930 | Rv1359 | Rv1359 | 3.7057528 |
| 931 | Rv3077 | Rv3077 | 3.705175 |
| 932 | Rv3685c | cyp137 | 3.7008438 |
| 933 | Rv3263 | Rv3263 | 3.6995103 |
| 934 | Rv1245c | Rv1245c | 3.6975727 |
| 935 | Rv3544c | fadE28 | 3.6885269 |
| 936 | Rv2805 | Rv2805 | 3.6866808 |
| 937 | Rv0911 | Rv0911 | 3.6842184 |
| 938 | Rv3546 | fadA5 | 3.6829252 |
| 939 | Rv2383c | mbtB | 3.6756928 |
| 940 | Rv3562 | fadE31 | 3.674732 |
| 941 | Rv0523c | Rv0523c | 3.6725638 |
| 942 | Rv3405c | Rv3405c | 3.670702 |
| 943 | Rv2159c | Rv2159c | 3.6634812 |
| 944 | Rv0942 | Rv0942 | 3.6595416 |
| 945 | Rv1011 | ispE | 3.657557 |
| 946 | Rv1800 | PPE28 | 3.6536117 |
| 947 | Rv2857c | Rv2857c | 3.6489131 |
| 948 | Rv0658c | Rv0658c | 3.637092 |
| 949 | Rv0944 | Rv0944 | 3.6365275 |
| 950 | Rv1518 | Rv1518 | 3.6326263 |
| 951 | Rv1689 | tyrS | 3.6297305 |
| 952 | Rv0454 | Rv0454 | 3.628013 |
| 953 | Rv0736 | rslA | 3.6232584 |
| 954 | Rv0368c | Rv0368c | 3.621969 |
| 955 | Rv3650 | PE33 | 3.6048586 |
| 956 | Rv2976c | ung | 3.6003873 |
| 957 | Rv3261 | fbiA | 3.5945551 |
| 958 | Rv1560 | vapB11 | 3.5926468 |
| 959 | Rv1530 | adh | 3.5828714 |
| 960 | Rv2160A | Rv2160A | 3.576596 |
| 961 | Rv3727 | Rv3727 | 3.5756617 |
| 962 | Rv2802c | Rv2802c | 3.5739975 |
| 963 | Rv2924c | fpg | 3.572603 |
| 964 | Rv0302 | Rv0302 | 3.5700905 |
| 965 | Rv2742c | Rv2742c | 3.5623329 |
| 966 | Rv1524 | Rv1524 | 3.5556018 |
| 967 | Rv3310 | sapM | 3.5550578 |
| 968 | Rv2319c | Rv2319c | 3.5548644 |
| 969 | Rv0146 | Rv0146 | 3.5541582 |
| 970 | Rv0892 | Rv0892 | 3.5501282 |
| 971 | Rv1495 | mazF4 | 3.5497415 |
| 972 | Rv2568c | Rv2568c | 3.5496495 |
| 973 | Rv0816c | thiX | 3.5491366 |
| 974 | Rv2831 | echA16 | 3.5477226 |
| 975 | Rv2211c | gcvT | 3.5460823 |
| 976 | Rv0052 | Rv0052 | 3.5431464 |
| 977 | Rv0033 | acpA | 3.5389042 |
| 978 | Rv2077c | Rv2077c | 3.5348458 |
| 979 | Rv1175c | fadH | 3.5300746 |
| 980 | Rv2592c | ruvB | 3.5284638 |
| 981 | Rv3312A | Rv3312A | 3.5217261 |
| 982 | Rv0648 | Rv0648 | 3.5212905 |
| 983 | Rv3725 | Rv3725 | 3.518507 |
| 984 | MTB000032 | argW | 3.5179734 |
| 985 | Rv3430c | Rv3430c | 3.5016603 |
| 986 | Rv3628 | ppa | 3.497686 |
| 987 | Rv0625c | Rv0625c | 3.4964106 |
| 988 | Rv2322c | rocD1 | 3.4894605 |
| 989 | Rv1259 | udgB | 3.484161 |
| 990 | Rv2569c | Rv2569c | 3.4818788 |
| 991 | Rv0320 | Rv0320 | 3.4759288 |
| 992 | Rv1538c | ansA | 3.4663186 |
| 993 | Rv2323c | Rv2323c | 3.4616692 |
| 994 | Rv0331 | Rv0331 | 3.4571342 |
| 995 | Rv1806 | PE20 | 3.4562142 |
| 996 | Rv1053c | Rv1053c | 3.4493768 |
| 997 | Rv0897c | Rv0897c | 3.446222 |
| 998 | Rv3247c | tmk | 3.440129 |
| 999 | Rv2517c | Rv2517c | 3.4389482 |
| 1000 | Rv0470A | Rv0470A | 3.428664 |
| 1001 | Rv3296 | lhr | 3.4084454 |
| 1002 | Rv1691 | Rv1691 | 3.4030414 |
| 1003 | Rv0089 | Rv0089 | 3.4026933 |
| 1004 | Rv3071 | Rv3071 | 3.4010775 |
| 1005 | Rv2896c | Rv2896c | 3.4003813 |
| 1006 | Rv3525c | Rv3525c | 3.3996298 |
| 1007 | Rv3308 | pmmB | 3.3975399 |
| 1008 | Rv1128c | Rv1128c | 3.397456 |
| 1009 | Rv0026 | Rv0026 | 3.3943176 |
| 1010 | Rv2069 | sigC | 3.3913295 |
| 1011 | Rv3413c | Rv3413c | 3.3852549 |
| 1012 | Rv0832 | PE_PGRS12 | 3.3795085 |
| 1013 | Rv3163c | Rv3163c | 3.375886 |
| 1014 | Rv0881 | Rv0881 | 3.3676696 |
| 1015 | Rv1126c | Rv1126c | 3.3629873 |
| 1016 | Rv2545 | vapB18 | 3.3594518 |
| 1017 | Rv1937 | Rv1937 | 3.3580408 |
| 1018 | Rv2382c | mbtC | 3.3547893 |
| 1019 | Rv0050 | ponA1 | 3.3507743 |
| 1020 | Rv0303 | Rv0303 | 3.348434 |
| 1021 | Rv3268 | Rv3268 | 3.3444767 |
| 1022 | Rv3202c | Rv3202c | 3.3363402 |
| 1023 | Rv2958c | Rv2958c | 3.3238177 |
| 1024 | Rv2226 | Rv2226 | 3.3221767 |
| 1025 | Rv1749c | Rv1749c | 3.3182538 |
| 1026 | Rv2874 | dipZ | 3.3174453 |
| 1027 | Rv2401A | Rv2401A | 3.3118098 |
| 1028 | Rv3496c | mce4D | 3.30591 |
| 1029 | Rv3313c | add | 3.2979076 |
| 1030 | Rv0746 | PE_PGRS9 | 3.2963254 |
| 1031 | Rv1840c | PE_PGRS34 | 3.2956367 |
| 1032 | Rv1959c | parE1 | 3.2826884 |
| 1033 | Rv3339c | icd1 | 3.2807586 |
| 1034 | Rv1151c | Rv1151c | 3.2801654 |
| 1035 | Rv0132c | fgd2 | 3.276 |
| 1036 | Rv1439c | Rv1439c | 3.2723882 |
| 1037 | Rv2192c | trpD | 3.2707098 |
| 1038 | Rv3661 | Rv3661 | 3.2703867 |
| 1039 | Rv3336c | trpS | 3.2676365 |
| 1040 | Rv0927c | Rv0927c | 3.2634935 |
| 1041 | Rv0340 | Rv0340 | 3.2620041 |
| 1042 | Rv0136 | cyp138 | 3.2614007 |
| 1043 | Rv2475c | Rv2475c | 3.2589788 |
| 1044 | Rv2290 | lppO | 3.2587702 |
| 1045 | Rv2288 | Rv2288 | 3.252317 |
| 1046 | Rv1137c | Rv1137c | 3.2315304 |
| 1047 | Rv3041c | Rv3041c | 3.2244873 |
| 1048 | Rv3669 | Rv3669 | 3.2188349 |
| 1049 | Rv2148c | Rv2148c | 3.2181344 |
| 1050 | Rv2551c | Rv2551c | 3.2146251 |
| 1051 | Rv2749 | Rv2749 | 3.2000277 |
| 1052 | Rv0212c | nadR | 3.1979074 |
| 1053 | Rv3111 | moaC1 | 3.196379 |
| 1054 | Rv3625c | mesJ | 3.1953633 |
| 1055 | Rv0039c | Rv0039c | 3.1938152 |
| 1056 | Rv0499 | Rv0499 | 3.1887739 |
| 1057 | Rv1621c | cydD | 3.1772747 |
| 1058 | Rv3861 | Rv3861 | 3.1731968 |
| 1059 | Rv1718 | Rv1718 | 3.1712976 |
| 1060 | Rv2832c | ugpC | 3.167704 |
| 1061 | Rv0193c | Rv0193c | 3.1597133 |
| 1062 | Rv3097c | lipY | 3.1550932 |
| 1063 | Rv0852 | fadD16 | 3.1531692 |
| 1064 | Rv1529 | fadD24 | 3.1503365 |
| 1065 | Rv1750c | fadD1 | 3.1472256 |
| 1066 | Rv3350c | PPE56 | 3.1384227 |
| 1067 | Rv3656c | Rv3656c | 3.134802 |
| 1068 | Rv2307D | Rv2307D | 3.125656 |
| 1069 | Rv3589 | mutY | 3.1253371 |
| 1070 | Rv2894c | xerC | 3.119659 |
| 1071 | Rv0028 | Rv0028 | 3.1118538 |
| 1072 | Rv0134 | ephF | 3.1118395 |
| 1073 | Rv3835 | Rv3835 | 3.1072037 |
| 1074 | Rv2234 | ptpA | 3.1054878 |
| 1075 | Rv2844 | Rv2844 | 3.1028075 |
| 1076 | Rv3107c | agpS | 3.0986211 |
| 1077 | Rv3553 | Rv3553 | 3.0929577 |
| 1078 | Rv3770B | Rv3770B | 3.0887063 |
| 1079 | Rv3175 | Rv3175 | 3.087671 |
| 1080 | Rv3393 | iunH | 3.0840178 |
| 1081 | Rv0802c | Rv0802c | 3.0779195 |
| 1082 | Rv1949c | Rv1949c | 3.063182 |
| 1083 | Rv3621c | PPE65 | 3.0527148 |
| 1084 | Rv3227 | aroA | 3.052677 |
| 1085 | Rv3069 | Rv3069 | 3.05239 |
| 1086 | Rv3796 | Rv3796 | 3.0515184 |
| 1087 | Rv1139c | Rv1139c | 3.0482807 |
| 1088 | Rv1815 | Rv1815 | 3.0468247 |
| 1089 | Rv3645 | Rv3645 | 3.0362892 |
| 1090 | Rv2746c | pgsA3 | 3.0359251 |
| 1091 | Rv1125 | Rv1125 | 3.0342567 |
| 1092 | Rv3549c | Rv3549c | 3.0319228 |
| 1093 | Rv0845 | Rv0845 | 3.0308821 |
| 1094 | Rv1081c | Rv1081c | 3.0243227 |
| 1095 | Rv0307c | Rv0307c | 3.0242097 |
| 1096 | Rv1012 | Rv1012 | 3.0203156 |
| 1097 | Rv1264 | Rv1264 | 3.0202575 |
| 1098 | Rv0768 | aldA | 3.0190134 |
| 1099 | Rv0804 | Rv0804 | 3.0173917 |
| 1100 | Rv1991c | mazF6 | 3.0086386 |
| 1101 | Rv3560c | fadE30 | 3.006252 |
| 1102 | Rv1877 | Rv1877 | 3.000603 |
| 1103 | Rv3599c | Rv3599c | 2.9998014 |
| 1104 | Rv3014c | ligA | 2.9966867 |
| 1105 | Rv2328 | PE23 | 2.9948907 |
| 1106 | Rv3572 | Rv3572 | 2.9935195 |
| 1107 | Rv0326 | Rv0326 | 2.9923563 |
| 1108 | Rv2385 | mbtJ | 2.9909449 |
| 1109 | MTB000009 | thrV | 2.9877214 |
| 1110 | Rv3918c | parA | 2.9843783 |
| 1111 | Rv2504c | scoA | 2.98308 |
| 1112 | Rv0583c | lpqN | 2.9815636 |
| 1113 | Rv1692 | Rv1692 | 2.9774334 |
| 1114 | Rv2027c | dosT | 2.9697518 |
| 1115 | Rv1345 | mbtM | 2.9671667 |
| 1116 | Rv3591c | Rv3591c | 2.9659297 |
| 1117 | Rv2171 | lppM | 2.9569235 |
| 1118 | Rv3728 | Rv3728 | 2.9565225 |
| 1119 | Rv2899c | fdhD | 2.9554439 |
| 1120 | Rv0887c | Rv0887c | 2.9548135 |
| 1121 | Rv0770 | Rv0770 | 2.9532623 |
| 1122 | Rv0266c | oplA | 2.9488215 |
| 1123 | Rv1802 | PPE30 | 2.9455903 |
| 1124 | Rv0917 | betP | 2.9435751 |
| 1125 | Rv2739c | Rv2739c | 2.9390097 |
| 1126 | Rv0484c | Rv0484c | 2.934226 |
| 1127 | Rv3531c | Rv3531c | 2.9284306 |
| 1128 | Rv0203 | Rv0203 | 2.9279351 |
| 1129 | Rv0429c | def | 2.9256258 |
| 1130 | Rv3209 | Rv3209 | 2.9255548 |
| 1131 | Rv0325 | Rv0325 | 2.923178 |
| 1132 | Rv1675c | cmr | 2.91994 |
| 1133 | Rv0541c | Rv0541c | 2.9137273 |
| 1134 | Rv2370c | Rv2370c | 2.9111505 |
| 1135 | Rv1239c | corA | 2.909402 |
| 1136 | Rv0899 | ompA | 2.908354 |
| 1137 | Rv1777 | cyp144 | 2.8908775 |
| 1138 | Rv1104 | Rv1104 | 2.8807201 |
| 1139 | Rv3717 | Rv3717 | 2.8801117 |
| 1140 | Rv0839 | Rv0839 | 2.8761694 |
| 1141 | Rv3021c | PPE47 | 2.8754163 |
| 1142 | Rv3758c | proV | 2.8750508 |
| 1143 | Rv3420c | rimI | 2.8747587 |
| 1144 | Rv2641 | cadI | 2.872925 |
| 1145 | Rv0727c | fucA | 2.8683734 |
| 1146 | Rv2801c | mazF9 | 2.8668866 |
| 1147 | Rv0041 | leuS | 2.8668077 |
| 1148 | Rv1780 | Rv1780 | 2.8648777 |
| 1149 | Rv3748 | Rv3748 | 2.8579311 |
| 1150 | Rv1025 | Rv1025 | 2.85781 |
| 1151 | Rv1219c | Rv1219c | 2.8505409 |
| 1152 | Rv2960c | Rv2960c | 2.8457317 |
| 1153 | Rv2002 | fabG3 | 2.844454 |
| 1154 | Rv0417 | thiG | 2.8361056 |
| 1155 | Rv2421c | nadD | 2.8336983 |
| 1156 | Rv0124 | PE_PGRS2 | 2.833205 |
| 1157 | Rv0138 | Rv0138 | 2.8305492 |
| 1158 | Rv0585c | Rv0585c | 2.8273244 |
| 1159 | Rv0074 | Rv0074 | 2.8245723 |
| 1160 | Rv2689c | Rv2689c | 2.8244505 |
| 1161 | Rv0901 | Rv0901 | 2.8243322 |
| 1162 | Rv1776c | Rv1776c | 2.8182466 |
| 1163 | Rv3770c | Rv3770c | 2.816438 |
| 1164 | Rv0653c | Rv0653c | 2.8142898 |
| 1165 | Rv3367 | PE_PGRS51 | 2.8119028 |
| 1166 | Rv0573c | pncB2 | 2.8112063 |
| 1167 | Rv2100 | Rv2100 | 2.8086956 |
| 1168 | Rv3360 | Rv3360 | 2.798274 |
| 1169 | Rv3634c | galE1 | 2.7949026 |
| 1170 | Rv3782 | glfT1 | 2.792114 |
| 1171 | Rv1371 | Rv1371 | 2.7894866 |
| 1172 | Rv2770c | PPE44 | 2.7886043 |
| 1173 | Rv0926c | Rv0926c | 2.7833385 |
| 1174 | Rv0989c | grcC2 | 2.7807856 |
| 1175 | Rv3885c | eccE2 | 2.778808 |
| 1176 | Rv3020c | esxS | 2.7758965 |
| 1177 | Rv0610c | Rv0610c | 2.7756042 |
| 1178 | Rv2062c | cobN | 2.771651 |
| 1179 | Rv1878 | glnA3 | 2.7660427 |
| 1180 | Rv3611 | Rv3611 | 2.765964 |
| 1181 | Rv3110 | moaB1 | 2.7635715 |
| 1182 | Rv3671c | Rv3671c | 2.75855 |
| 1183 | Rv1817 | Rv1817 | 2.7538855 |
| 1184 | Rv2834c | ugpE | 2.753661 |
| 1185 | Rv0880 | Rv0880 | 2.7474318 |
| 1186 | Rv2074 | Rv2074 | 2.747232 |
| 1187 | Rv1243c | PE_PGRS23 | 2.7458146 |
| 1188 | Rv1170 | mshB | 2.7457108 |
| 1189 | Rv0398c | Rv0398c | 2.7427256 |
| 1190 | Rv2700 | Rv2700 | 2.740716 |
| 1191 | Rv2666 | Rv2666 | 2.7371788 |
| 1192 | Rv0413 | mutT3 | 2.735588 |
| 1193 | Rv3644c | Rv3644c | 2.7314258 |
| 1194 | Rv3575c | Rv3575c | 2.7247298 |
| 1195 | Rv0758 | phoR | 2.7245417 |
| 1196 | Rv3297 | nei | 2.7117414 |
| 1197 | Rv3578 | arsB2 | 2.7108707 |
| 1198 | MTB000072 | mcr10 | 2.7051673 |
| 1199 | Rv0995 | rimJ | 2.700624 |
| 1200 | Rv2679 | echA15 | 2.697998 |
| 1201 | Rv3270 | ctpC | 2.6976793 |
| 1202 | Rv0367c | Rv0367c | 2.6970704 |
| 1203 | Rv3754 | tyrA | 2.6965375 |
| 1204 | Rv3886c | mycP2 | 2.6937342 |
| 1205 | Rv2701c | suhB | 2.6931362 |
| 1206 | Rv1907c | Rv1907c | 2.6848319 |
| 1207 | Rv0520 | Rv0520 | 2.678297 |
| 1208 | Rv1147 | Rv1147 | 2.6781242 |
| 1209 | Rv3666c | dppA | 2.674516 |
| 1210 | Rv1402 | priA | 2.6741045 |
| 1211 | Rv0035 | fadD34 | 2.6740065 |
| 1212 | Rv0374c | Rv0374c | 2.6728 |
| 1213 | Rv3047c | Rv3047c | 2.6689985 |
| 1214 | Rv3450c | eccB4 | 2.6686852 |
| 1215 | Rv1180 | pks3 | 2.6659184 |
| 1216 | Rv3787c | Rv3787c | 2.6642478 |
| 1217 | Rv0463 | Rv0463 | 2.6586611 |
| 1218 | Rv2761c | hsdS | 2.6557953 |
| 1219 | Rv2622 | Rv2622 | 2.6491392 |
| 1220 | Rv3794 | embA | 2.647172 |
| 1221 | Rv2821c | Rv2821c | 2.6470726 |
| 1222 | Rv0347 | Rv0347 | 2.6458015 |
| 1223 | Rv3785 | Rv3785 | 2.6376917 |
| 1224 | Rv2153c | murG | 2.6364238 |
| 1225 | Rv2985 | mutT1 | 2.635734 |
| 1226 | Rv3739c | PPE67 | 2.6351635 |
| 1227 | Rv1773c | Rv1773c | 2.630098 |
| 1228 | Rv2141c | Rv2141c | 2.627849 |
| 1229 | Rv0017c | rodA | 2.622775 |
| 1230 | Rv0471c | Rv0471c | 2.6159506 |
| 1231 | Rv2624c | Rv2624c | 2.612966 |
| 1232 | Rv0565c | Rv0565c | 2.6120734 |
| 1233 | Rv3016 | lpqA | 2.6117818 |
| 1234 | Rv3522 | ltp4 | 2.6084146 |
| 1235 | Rv0609A | Rv0609A | 2.6064293 |
| 1236 | Rv3183 | Rv3183 | 2.6056988 |
| 1237 | Rv1141c | echA11 | 2.6012545 |
| 1238 | Rv2318 | uspC | 2.5961027 |
| 1239 | Rv0116c | ldtA | 2.59569 |
| 1240 | Rv2528c | mrr | 2.5942593 |
| 1241 | Rv2866 | relG | 2.5926926 |
| 1242 | Rv0669c | Rv0669c | 2.585966 |
| 1243 | Rv3078 | hab | 2.5852685 |
| 1244 | Rv3025c | iscS | 2.584697 |
| 1245 | Rv3792 | aftA | 2.5827792 |
| 1246 | Rv0453 | PPE11 | 2.5818584 |
| 1247 | Rv3688c | Rv3688c | 2.581588 |
| 1248 | Rv2820c | Rv2820c | 2.578642 |
| 1249 | Rv2905 | lppW | 2.5742319 |
| 1250 | Rv3535c | hsaG | 2.5731382 |
| 1251 | Rv1753c | PPE24 | 2.5713542 |
| 1252 | Rv2885c | Rv2885c | 2.5684118 |
| 1253 | Rv3358 | relK | 2.5672908 |
| 1254 | Rv1525 | wbbL2 | 2.5663803 |
| 1255 | Rv0525 | Rv0525 | 2.5641422 |
| 1256 | Rv1512 | epiA | 2.5565073 |
| 1257 | Rv1040c | PE8 | 2.5470417 |
| 1258 | Rv2197c | Rv2197c | 2.5374024 |
| 1259 | Rv1857 | modA | 2.534454 |
| 1260 | Rv3540c | ltp2 | 2.5307133 |
| 1261 | Rv2413c | Rv2413c | 2.5282583 |
| 1262 | Rv1992c | ctpG | 2.5265563 |
| 1263 | Rv3113 | Rv3113 | 2.5221372 |
| 1264 | Rv2830c | vapB22 | 2.5221362 |
| 1265 | Rv2918c | glnD | 2.521925 |
| 1266 | Rv0726c | Rv0726c | 2.515353 |
| 1267 | Rv3037c | Rv3037c | 2.5135093 |
| 1268 | Rv3790 | dprE1 | 2.5117712 |
| 1269 | Rv3705c | Rv3705c | 2.5099797 |
| 1270 | Rv0420c | Rv0420c | 2.5035117 |
| 1271 | Rv3476c | kgtP | 2.4948604 |
| 1272 | Rv1527c | pks5 | 2.494598 |
| 1273 | Rv3579c | Rv3579c | 2.4935617 |
| 1274 | Rv0578c | PE_PGRS7 | 2.4853647 |
| 1275 | Rv1650 | pheT | 2.4843557 |
| 1276 | Rv1788 | PE18 | 2.4791527 |
| 1277 | Rv1367c | Rv1367c | 2.4790378 |
| 1278 | Rv2414c | Rv2414c | 2.4763174 |
| 1279 | Rv2764c | thyA | 2.4763014 |
| 1280 | Rv1429 | Rv1429 | 2.4737084 |
| 1281 | Rv1424c | Rv1424c | 2.4555638 |
| 1282 | Rv2394 | ggtB | 2.4507601 |
| 1283 | Rv0800 | pepC | 2.4490166 |
| 1284 | Rv1373 | Rv1373 | 2.4476576 |
| 1285 | Rv2259 | mscR | 2.4464085 |
| 1286 | Rv3789 | Rv3789 | 2.4424512 |
| 1287 | Rv3309c | upp | 2.4387043 |
| 1288 | Rv2712c | Rv2712c | 2.436856 |
| 1289 | Rv1696 | recN | 2.4336014 |
| 1290 | Rv1176c | Rv1176c | 2.431201 |
| 1291 | Rv2070c | cobK | 2.4295094 |
| 1292 | Rv3898c | Rv3898c | 2.4271164 |
| 1293 | Rv0186 | bglS | 2.424084 |
| 1294 | Rv0997 | Rv0997 | 2.417144 |
| 1295 | Rv0893c | Rv0893c | 2.4144392 |
| 1296 | Rv3732 | Rv3732 | 2.4137592 |
| 1297 | Rv2678c | hemE | 2.4060032 |
| 1298 | Rv2539c | aroK | 2.4024436 |
| 1299 | Rv2665 | Rv2665 | 2.4005609 |
| 1300 | Rv0579 | Rv0579 | 2.3968768 |
| 1301 | Rv0238 | Rv0238 | 2.3932722 |
| 1302 | MTB000046 | serV | 2.3817728 |
| 1303 | Rv1918c | PPE35 | 2.3807976 |
| 1304 | Rv3312c | Rv3312c | 2.3763313 |
| 1305 | Rv3253c | Rv3253c | 2.3755317 |
| 1306 | Rv1607 | chaA | 2.3637903 |
| 1307 | Rv0743c | Rv0743c | 2.362261 |
| 1308 | Rv2695 | Rv2695 | 2.3591757 |
| 1309 | Rv2403c | lppR | 2.3557594 |
| 1310 | Rv0778 | cyp126 | 2.3524878 |
| 1311 | Rv0128 | Rv0128 | 2.3509932 |
| 1312 | Rv1682 | Rv1682 | 2.3509083 |
| 1313 | Rv2026c | Rv2026c | 2.3498824 |
| 1314 | Rv3677c | Rv3677c | 2.3441527 |
| 1315 | Rv0553 | menC | 2.3401852 |
| 1316 | Rv2585c | Rv2585c | 2.3386168 |
| 1317 | Rv1850 | ureC | 2.3383298 |
| 1318 | Rv2616 | Rv2616 | 2.331881 |
| 1319 | Rv2549c | vapC20 | 2.3313339 |
| 1320 | Rv2511 | orn | 2.3199177 |
| 1321 | Rv2570 | Rv2570 | 2.3115904 |
| 1322 | Rv0401 | Rv0401 | 2.310299 |
| 1323 | Rv1343c | lprD | 2.308556 |
| 1324 | Rv0534c | menA | 2.3050845 |
| 1325 | Rv3487c | lipF | 2.3020241 |
| 1326 | Rv2911 | dacB2 | 2.3000069 |
| 1327 | Rv3481c | Rv3481c | 2.2995162 |
| 1328 | Rv3125c | PPE49 | 2.2982006 |
| 1329 | Rv0406c | Rv0406c | 2.2980785 |
| 1330 | Rv0729 | xylB | 2.2878864 |
| 1331 | Rv3556c | fadA6 | 2.2764318 |
| 1332 | Rv0131c | fadE1 | 2.2711747 |
| 1333 | Rv2401 | Rv2401 | 2.2704906 |
| 1334 | Rv2738c | Rv2738c | 2.2639253 |
| 1335 | Rv3137 | Rv3137 | 2.26072 |
| 1336 | Rv0599c | vapB27 | 2.2583976 |
| 1337 | Rv3781 | rfbE | 2.257428 |
| 1338 | Rv2298 | Rv2298 | 2.2550375 |
| 1339 | Rv1076 | lipU | 2.243782 |
| 1340 | Rv2669 | Rv2669 | 2.2409248 |
| 1341 | Rv3234c | tgs3 | 2.2387838 |
| 1342 | Rv1251c | Rv1251c | 2.2343495 |
| 1343 | Rv1567c | Rv1567c | 2.2226915 |
| 1344 | Rv1745c | idi | 2.2075195 |
| 1345 | Rv2541 | Rv2541 | 2.179051 |
| 1346 | Rv3761c | fadE36 | 2.175231 |
| 1347 | Rv1032c | trcS | 2.1730857 |
| 1348 | Rv1121 | zwf1 | 2.166517 |
| 1349 | Rv0438c | moeA2 | 2.1651442 |
| 1350 | Rv3176c | mesT | 2.1642814 |
| 1351 | Rv2514c | Rv2514c | 2.1566637 |
| 1352 | MTB000020 | rrl | 2.142046 |
| 1353 | Rv2164c | Rv2164c | 2.1395853 |
| 1354 | Rv1922 | Rv1922 | 2.13311 |
| 1355 | Rv3888c | Rv3888c | 2.132445 |
| 1356 | Rv3242c | Rv3242c | 2.1271355 |
| 1357 | Rv1071c | echA9 | 2.1255639 |
| 1358 | Rv0335c | PE6 | 2.1008446 |
| 1359 | Rv1981c | nrdF1 | 2.0991945 |
| 1360 | Rv0318c | Rv0318c | 2.0974483 |
| 1361 | Rv3404c | Rv3404c | 2.0585876 |
| 1362 | Rv2613c | Rv2613c | 2.0487578 |
| 1363 | Rv0443 | Rv0443 | 2.0400243 |
| 1364 | Rv0382c | pyrE | 2.0369337 |
| 1365 | Rv1890c | Rv1890c | 2.0067658 |
