## Supplementary Table 3 for "Diagnostic utility of in vivo expressed mycobacterial RNA transcripts in pleural fluid for the differential diagnosis of tuberculous pleuritis"

**ANNEXURE II**

**DOWNREGULATED GENES IN BIOPSY SAMPLES OF TB PLEURITIS PATIENTS**

| **S.No.** | **Gene name** | **Gene ID** | **FC ([test] Vs [control])** |
| --- | --- | --- | --- |
| 1 | MTB000048 | argU | -292.89496 |
| 2 | MTB000034 | lysU | -225.94174 |
| 3 | MTB000035 | valT | -198.80438 |
| 4 | MTB000033 | hisT | -198.25752 |
| 5 | MTB000067 | mcr7 | -112.92638 |
| 6 | Rv3874 | esxB | -95.74769 |
| 7 | Rv1792 | esxM | -88.23279 |
| 8 | Rv2780 | ald | -86.5684 |
| 9 | Rv1738 | Rv1738 | -85.42212 |
| 10 | Rv3875 | esxA | -80.901535 |
| 11 | Rv1871c | Rv1871c | -67.97308 |
| 12 | Rv1038c | esxJ | -62.227146 |
| 13 | Rv3615c | espC | -59.441936 |
| 14 | Rv0288 | esxH | -59.37261 |
| 15 | Rv2031c | hspX | -49.428497 |
| 16 | Rv1794 | Rv1794 | -49.10399 |
| 17 | Rv3131 | Rv3131 | -47.374367 |
| 18 | Rv2005c | Rv2005c | -43.961494 |
| 19 | Rv1793 | esxN | -42.27542 |
| 20 | Rv0287 | esxG | -41.081215 |
| 21 | Rv3620c | esxW | -37.700584 |
| 22 | Rv3130c | tgs1 | -33.592316 |
| 23 | Rv3456c | rplQ | -33.513344 |
| 24 | Rv2441c | rpmA | -31.810545 |
| 25 | Rv2840c | Rv2840c | -30.983717 |
| 26 | Rv2346c | esxO | -30.531925 |
| 27 | Rv3219 | whiB1 | -29.31121 |
| 28 | Rv2347c | esxP | -28.47694 |
| 29 | Rv0108c | Rv0108c | -28.33726 |
| 30 | Rv3841 | bfrB | -26.4624 |
| 31 | Rv3461c | rpmJ | -26.167114 |
| 32 | Rv1986 | Rv1986 | -25.717705 |
| 33 | Rv0569 | Rv0569 | -25.381556 |
| 34 | Rv1072 | Rv1072 | -23.942217 |
| 35 | Rv1197 | esxK | -23.555288 |
| 36 | Rv1198 | esxL | -23.268957 |
| 37 | Rv3246c | mtrA | -21.973185 |
| 38 | Rv3583c | Rv3583c | -20.969107 |
| 39 | Rv1571 | Rv1571 | -20.165783 |
| 40 | Rv2628 | Rv2628 | -20.119833 |
| 41 | Rv2632c | Rv2632c | -19.975979 |
| 42 | Rv2623 | TB31.7 | -19.238316 |
| 43 | Rv0683 | rpsG | -18.507746 |
| 44 | MTB000018 | argV | -18.272331 |
| 45 | Rv0280 | PPE3 | -17.315552 |
| 46 | Rv2348c | Rv2348c | -16.772907 |
| 47 | Rv0667 | rpoB | -16.36804 |
| 48 | Rv2745c | clgR | -16.031988 |
| 49 | Rv3872 | PE35 | -16.026373 |
| 50 | Rv3127 | Rv3127 | -15.575929 |
| 51 | Rv2558 | Rv2558 | -15.264497 |
| 52 | Rv3134c | Rv3134c | -15.23047 |
| 53 | Rv3870 | eccCa1 | -14.939124 |
| 54 | Rv2676c | Rv2676c | -14.809266 |
| 55 | Rv3763 | lpqH | -14.506678 |
| 56 | Rv2007c | fdxA | -14.316458 |
| 57 | Rv3052c | nrdI | -14.243311 |
| 58 | Rv3619c | esxV | -14.086879 |
| 59 | Rv1653 | argJ | -13.4230995 |
| 60 | Rv1996 | Rv1996 | -13.3779955 |
| 61 | Rv2627c | Rv2627c | -13.324351 |
| 62 | Rv2302 | Rv2302 | -12.911647 |
| 63 | Rv1466 | Rv1466 | -12.854985 |
| 64 | Rv2708c | Rv2708c | -12.640822 |
| 65 | Rv3847 | Rv3847 | -12.384436 |
| 66 | Rv0709 | rpmC | -12.1819 |
| 67 | Rv3133c | devR | -12.065961 |
| 68 | Rv0285 | PE5 | -12.061814 |
| 69 | Rv3873 | PPE68 | -11.97426 |
| 70 | Rv0079 | Rv0079 | -11.700927 |
| 71 | Rv0286 | PPE4 | -11.618487 |
| 72 | Rv2442c | rplU | -11.5909 |
| 73 | Rv3864 | espE | -11.52809 |
| 74 | Rv3865 | espF | -11.43716 |
| 75 | Rv3053c | nrdH | -11.425241 |
| 76 | Rv3418c | groES | -11.415215 |
| 77 | Rv2032 | acg | -11.400682 |
| 78 | Rv3612c | Rv3612c | -11.147221 |
| 79 | Rv0054 | ssb | -11.143642 |
| 80 | Rv3871 | eccCb1 | -10.735114 |
| 81 | Rv1333 | Rv1333 | -10.696857 |
| 82 | Rv3648c | cspA | -10.651975 |
| 83 | Rv2694c | Rv2694c | -10.531309 |
| 84 | Rv3289c | Rv3289c | -10.47626 |
| 85 | Rv0833 | PE_PGRS13 | -10.444836 |
| 86 | Rv2925c | rnc | -10.410289 |
| 87 | Rv1261c | Rv1261c | -10.40213 |
| 88 | Rv0824c | desA1 | -10.34934 |
| 89 | Rv0080 | Rv0080 | -10.295157 |
| 90 | Rv1791 | PE19 | -10.250249 |
| 91 | Rv0220 | lipC | -10.160612 |
| 92 | Rv0188 | Rv0188 | -10.078237 |
| 93 | Rv2030c | Rv2030c | -10.003398 |
| 94 | Rv1211 | Rv1211 | -9.994364 |
| 95 | Rv1322A | Rv1322A | -9.740497 |
| 96 | Rv3229c | desA3 | -9.7045355 |
| 97 | Rv3614c | espD | -9.689875 |
| 98 | Rv0682 | rpsL | -9.675449 |
| 99 | Rv0289 | espG3 | -9.673506 |
| 100 | Rv2527 | vapC17 | -9.614825 |
| 101 | Rv2110c | prcB | -9.55364 |
| 102 | Rv0685 | tuf | -9.457506 |
| 103 | Rv0207c | Rv0207c | -9.40268 |
| 104 | Rv3913 | trxB2 | -9.345115 |
| 105 | Rv2626c | hrp1 | -9.26448 |
| 106 | Rv2557 | Rv2557 | -9.2046995 |
| 107 | Rv3679 | Rv3679 | -9.123982 |
| 108 | Rv3829c | Rv3829c | -9.054577 |
| 109 | Rv1827 | garA | -8.871481 |
| 110 | Rv2526 | vapB17 | -8.870557 |
| 111 | Rv3613c | Rv3613c | -8.835034 |
| 112 | Rv3051c | nrdE | -8.819512 |
| 113 | Rv2986c | hupB | -8.593661 |
| 114 | Rv3827c | Rv3827c | -8.383067 |
| 115 | Rv3412 | Rv3412 | -8.376018 |
| 116 | Rv0639 | nusG | -8.367222 |
| 117 | Rv0349 | Rv0349 | -8.33934 |
| 118 | MTB000022 | leuW | -8.2599745 |
| 119 | Rv2182c | Rv2182c | -8.231682 |
| 120 | Rv0056 | rplI | -8.149177 |
| 121 | Rv3859c | gltB | -7.823244 |
| 122 | Rv0708 | rplP | -7.8009586 |
| 123 | Rv3221A | rshA | -7.7694454 |
| 124 | Rv0485 | Rv0485 | -7.550214 |
| 125 | Rv1109c | Rv1109c | -7.5328774 |
| 126 | Rv1398c | vapB10 | -7.5174794 |
| 127 | Rv1073 | Rv1073 | -7.418421 |
| 128 | Rv2583c | relA | -7.3825684 |
| 129 | Rv2904c | rplS | -7.3537927 |
| 130 | Rv2050 | Rv2050 | -7.325852 |
| 131 | Rv3922c | Rv3922c | -7.2734385 |
| 132 | Rv0516c | Rv0516c | -7.201276 |
| 133 | Rv1880c | cyp140 | -7.159095 |
| 134 | Rv0948c | Rv0948c | -7.139301 |
| 135 | Rv1908c | katG | -7.124919 |
| 136 | Rv3891c | esxD | -7.1173096 |
| 137 | Rv0634B | rpmG2 | -7.100228 |
| 138 | Rv3001c | ilvC | -7.0193453 |
| 139 | Rv1829 | Rv1829 | -6.97785 |
| 140 | Rv2457c | clpX | -6.956972 |
| 141 | Rv0088 | Rv0088 | -6.8773966 |
| 142 | Rv3734c | tgs2 | -6.7780676 |
| 143 | Rv0859 | fadA | -6.7141933 |
| 144 | Rv2245 | kasA | -6.6928625 |
| 145 | Rv2882c | frr | -6.679616 |
| 146 | Rv3290c | lat | -6.647167 |
| 147 | Rv2461c | clpP1 | -6.637783 |
| 148 | Rv2744c | 35kd_ag | -6.5842795 |
| 149 | MTB000069 | mpr12 | -6.580009 |
| 150 | Rv0166 | fadD5 | -6.5688553 |
| 151 | Rv0638 | secE1 | -6.543274 |
| 152 | Rv2740 | ephG | -6.5221 |
| 153 | Rv2004c | Rv2004c | -6.4782743 |
| 154 | Rv1174c | TB8.4 | -6.4622674 |
| 155 | Rv1461 | Rv1461 | -6.3857946 |
| 156 | Rv2699c | Rv2699c | -6.3249106 |
| 157 | Rv0475 | hbhA | -6.2661715 |
| 158 | Rv1795 | eccD5 | -6.226733 |
| 159 | Rv3867 | espH | -6.225882 |
| 160 | Rv0490 | senX3 | -6.2191114 |
| 161 | Rv2455c | Rv2455c | -6.215966 |
| 162 | Rv2204c | Rv2204c | -6.2086124 |
| 163 | Rv1305 | atpE | -6.175003 |
| 164 | Rv0503c | cmaA2 | -6.1577363 |
| 165 | Rv3846 | sodA | -6.1456943 |
| 166 | Rv2094c | tatA | -6.116465 |
| 167 | Rv3371 | Rv3371 | -6.1104455 |
| 168 | Rv0279c | PE_PGRS4 | -6.0645795 |
| 169 | Rv2930 | fadD26 | -6.047177 |
| 170 | Rv3462c | infA | -6.028695 |
| 171 | Rv2162c | PE_PGRS38 | -6.0234327 |
| 172 | Rv0297 | PE_PGRS5 | -5.9568324 |
| 173 | Rv3592 | mhuD | -5.9564605 |
| 174 | Rv1828 | Rv1828 | -5.9273868 |
| 175 | Rv2406c | Rv2406c | -5.9186387 |
| 176 | Rv1797 | eccE5 | -5.914644 |
| 177 | Rv3616c | espA | -5.8699427 |
| 178 | Rv0747 | PE_PGRS10 | -5.861481 |
| 179 | Rv1476 | Rv1476 | -5.8606772 |
| 180 | Rv3491 | Rv3491 | -5.8511543 |
| 181 | Rv0167 | yrbE1A | -5.8400893 |
| 182 | Rv3880c | espL | -5.7792892 |
| 183 | MTB000066 | mcr5 | -5.7638626 |
| 184 | Rv3849 | espR | -5.7442417 |
| 185 | Rv2875 | mpt70 | -5.6394053 |
| 186 | Rv0640 | rplK | -5.6218863 |
| 187 | Rv0106 | Rv0106 | -5.6060505 |
| 188 | Rv0491 | regX3 | -5.577426 |
| 189 | Rv1462 | Rv1462 | -5.561803 |
| 190 | Rv2244 | acpM | -5.537735 |
| 191 | Rv2778c | Rv2778c | -5.4999666 |
| 192 | Rv2916c | ffh | -5.4953976 |
| 193 | Rv0426c | Rv0426c | -5.4873314 |
| 194 | Rv0706 | rplV | -5.4815817 |
| 195 | Rv0896 | gltA2 | -5.47677 |
| 196 | Rv0572c | Rv0572c | -5.391855 |
| 197 | Rv3890c | esxC | -5.389527 |
| 198 | Rv3221c | TB7.3 | -5.381509 |
| 199 | Rv0502 | Rv0502 | -5.333399 |
| 200 | MTB000012 | aspT | -5.306906 |
| 201 | Rv2144c | Rv2144c | -5.2833543 |
| 202 | Rv2111c | pup | -5.2758064 |
| 203 | Rv2016 | Rv2016 | -5.275081 |
| 204 | Rv2629 | Rv2629 | -5.2597404 |
| 205 | Rv2200c | ctaC | -5.2144876 |
| 206 | Rv1487 | Rv1487 | -5.207114 |
| 207 | Rv3597c | lsr2 | -5.1925273 |
| 208 | Rv2336 | Rv2336 | -5.19073 |
| 209 | Rv3460c | rpsM | -5.190634 |
| 210 | Rv1309 | atpG | -5.121548 |
| 211 | Rv1489A | Rv1489A | -5.1047354 |
| 212 | Rv2428 | ahpC | -5.084965 |
| 213 | Rv2220 | glnA1 | -5.0656505 |
| 214 | Rv0239 | vapB24 | -5.060616 |
| 215 | Rv0055 | rpsR1 | -5.0545473 |
| 216 | Rv0145 | Rv0145 | -5.0543127 |
| 217 | Rv0247c | Rv0247c | -5.052578 |
| 218 | Rv0715 | rplX | -5.037733 |
| 219 | Rv2021c | Rv2021c | -5.036398 |
| 220 | Rv0500A | Rv0500A | -5.0362253 |
| 221 | Rv1843c | guaB1 | -5.0037227 |
| 222 | Rv0668 | rpoC | -4.9817166 |
| 223 | Rv0932c | pstS2 | -4.9665203 |
| 224 | Rv3207c | Rv3207c | -4.965321 |
| 225 | Rv0283 | eccB3 | -4.910467 |
| 226 | Rv0660c | mazE2 | -4.8566494 |
| 227 | Rv0937c | mku | -4.841739 |
| 228 | Rv0282 | eccA3 | -4.815292 |
| 229 | Rv0659c | mazF2 | -4.808278 |
| 230 | Rv1846c | blaI | -4.79011 |
| 231 | Rv2555c | alaS | -4.773761 |
| 232 | MTB000008 | trpT | -4.7729607 |
| 233 | Rv0078A | Rv0078A | -4.7678466 |
| 234 | Rv1293 | lysA | -4.732878 |
| 235 | Rv1883c | Rv1883c | -4.713829 |
| 236 | Rv0211 | pckA | -4.6993723 |
| 237 | Rv2218 | lipA | -4.692031 |
| 238 | Rv0829 | Rv0829 | -4.6706877 |
| 239 | Rv3301c | phoY1 | -4.648821 |
| 240 | Rv0292 | eccE3 | -4.6459417 |
| 241 | Rv1321 | Rv1321 | -4.6442986 |
| 242 | Rv2771c | Rv2771c | -4.6207256 |
| 243 | Rv0700 | rpsJ | -4.610342 |
| 244 | Rv1736c | narX | -4.608708 |
| 245 | Rv3144c | PPE52 | -4.591887 |
| 246 | Rv1783 | eccC5 | -4.5860066 |
| 247 | Rv1703c | Rv1703c | -4.5146704 |
| 248 | Rv0922 | Rv0922 | -4.511688 |
| 249 | Rv0103c | ctpB | -4.4967937 |
| 250 | Rv3733c | Rv3733c | -4.496471 |
| 251 | Rv2496c | bkdB | -4.48059 |
| 252 | Rv0826 | Rv0826 | -4.45518 |
| 253 | Rv1297 | rho | -4.4549413 |
| 254 | Rv1638A | Rv1638A | -4.4229636 |
| 255 | Rv3680 | Rv3680 | -4.42065 |
| 256 | Rv2625c | Rv2625c | -4.3834896 |
| 257 | Rv1658 | argG | -4.380278 |
| 258 | Rv1904 | Rv1904 | -4.3785586 |
| 259 | Rv0298 | Rv0298 | -4.360599 |
| 260 | Rv0500B | Rv0500B | -4.325384 |
| 261 | Rv3295 | Rv3295 | -4.309766 |
| 262 | Rv3389c | htdY | -4.304274 |
| 263 | Rv2112c | dop | -4.2536798 |
| 264 | Rv0157A | Rv0157A | -4.231027 |
| 265 | Rv2476c | gdh | -4.2148933 |
| 266 | Rv1352 | Rv1352 | -4.1934986 |
| 267 | Rv1799 | lppT | -4.1221895 |
| 268 | Rv1298 | rpmE | -4.119998 |
| 269 | Rv1479 | moxR1 | -4.1091633 |
| 270 | Rv1821 | secA2 | -4.1090536 |
| 271 | Rv3143 | Rv3143 | -4.103448 |
| 272 | Rv0176 | Rv0176 | -4.085548 |
| 273 | Rv2693c | Rv2693c | -4.0851035 |
| 274 | Rv1501 | Rv1501 | -4.0630226 |
| 275 | Rv1932 | tpx | -4.048323 |
| 276 | Rv1080c | greA | -4.0094337 |
| 277 | Rv1265 | Rv1265 | -4.0038896 |
| 278 | Rv3132c | devS | -3.9982855 |
| 279 | Rv1813c | Rv1813c | -3.964869 |
| 280 | Rv3241c | Rv3241c | -3.8981469 |
| 281 | Rv1159A | Rv1159A | -3.8820388 |
| 282 | Rv2996c | serA1 | -3.872894 |
| 283 | Rv1641 | infC | -3.8659475 |
| 284 | Rv3519 | Rv3519 | -3.8600323 |
| 285 | Rv1980c | mpt64 | -3.8484716 |
| 286 | Rv3596c | clpC1 | -3.847161 |
| 287 | Rv1342c | Rv1342c | -3.8466384 |
| 288 | Rv0313 | Rv0313 | -3.8403475 |
| 289 | MTB000010 | lysT | -3.8383517 |
| 290 | Rv0019c | fhaB | -3.8163533 |
| 291 | Rv3528c | Rv3528c | -3.8101444 |
| 292 | Rv3801c | fadD32 | -3.8084002 |
| 293 | Rv3881c | espB | -3.7913215 |
| 294 | Rv1152 | Rv1152 | -3.7893775 |
| 295 | Rv2982c | gpdA2 | -3.7738345 |
| 296 | Rv3411c | guaB2 | -3.7668622 |
| 297 | Rv0716 | rplE | -3.7494845 |
| 298 | Rv2454c | Rv2454c | -3.7390826 |
| 299 | Rv2431c | PE25 | -3.7333364 |
| 300 | Rv1654 | argB | -3.7219484 |
| 301 | Rv0425c | ctpH | -3.713178 |
| 302 | Rv2372c | Rv2372c | -3.69678 |
| 303 | Rv0637 | hadC | -3.6843157 |
| 304 | Rv3869 | eccB1 | -3.6732457 |
| 305 | Rv0281 | Rv0281 | -3.6636095 |
| 306 | Rv0724 | sppA | -3.6588554 |
| 307 | Rv0755c | PPE12 | -3.657488 |
| 308 | Rv1201c | dapD | -3.6466815 |
| 309 | Rv3828c | Rv3828c | -3.642027 |
| 310 | Rv1893 | Rv1893 | -3.6335843 |
| 311 | Rv0958 | Rv0958 | -3.630563 |
| 312 | Rv2187 | fadD15 | -3.6293817 |
| 313 | Rv3843c | Rv3843c | -3.5736187 |
| 314 | Rv3724B | cut5b | -3.5645237 |
| 315 | Rv0609 | vapC28 | -3.5594606 |
| 316 | Rv2029c | pfkB | -3.5448124 |
| 317 | Rv1782 | eccB5 | -3.5291996 |
| 318 | Rv1362c | Rv1362c | -3.5230837 |
| 319 | Rv3479 | Rv3479 | -3.5110905 |
| 320 | Rv2531c | Rv2531c | -3.5049827 |
| 321 | Rv1535 | Rv1535 | -3.4901276 |
| 322 | Rv1733c | Rv1733c | -3.4719021 |
| 323 | Rv3240c | secA1 | -3.4718113 |
| 324 | Rv3678c | Rv3678c | -3.4447985 |
| 325 | Rv2710 | sigB | -3.4335508 |
| 326 | Rv1872c | lldD2 | -3.3999407 |
| 327 | Rv2453c | mobA | -3.3993196 |
| 328 | Rv3831 | Rv3831 | -3.3965406 |
| 329 | Rv2879c | Rv2879c | -3.3750052 |
| 330 | Rv2169c | Rv2169c | -3.3606706 |
| 331 | Rv3155 | nuoK | -3.3508847 |
| 332 | Rv1444c | Rv1444c | -3.346249 |
| 333 | Rv1547 | dnaE1 | -3.3407278 |
| 334 | Rv1909c | furA | -3.328931 |
| 335 | Rv3164c | moxR3 | -3.3223548 |
| 336 | Rv0608 | vapB28 | -3.3145034 |
| 337 | Rv0831c | Rv0831c | -3.30035 |
| 338 | Rv2970A | Rv2970A | -3.2943733 |
| 339 | Rv3058c | Rv3058c | -3.2612147 |
| 340 | Rv3924c | rpmH | -3.2435575 |
| 341 | Rv0178 | Rv0178 | -3.2282865 |
| 342 | Rv2967c | pca | -3.2271354 |
| 343 | Rv0722 | rpmD | -3.2271087 |
| 344 | Rv1156 | Rv1156 | -3.2269204 |
| 345 | Rv3302c | glpD2 | -3.219474 |
| 346 | Rv1830 | Rv1830 | -3.2093613 |
| 347 | Rv0455c | Rv0455c | -3.1932487 |
| 348 | Rv1923 | lipD | -3.1914473 |
| 349 | Rv3206c | moeB1 | -3.1893368 |
| 350 | Rv0719 | rplF | -3.183394 |
| 351 | Rv3799c | accD4 | -3.1793091 |
| 352 | Rv0064 | Rv0064 | -3.1667402 |
| 353 | Rv2219 | Rv2219 | -3.1617918 |
| 354 | Rv0364 | Rv0364 | -3.1585193 |
| 355 | Rv0046c | ino1 | -3.1536708 |
| 356 | Rv0174 | mce1F | -3.1435888 |
| 357 | Rv2271 | Rv2271 | -3.1336262 |
| 358 | Rv3633 | Rv3633 | -3.1301434 |
| 359 | Rv2202c | adoK | -3.1245399 |
| 360 | Rv0991c | Rv0991c | -3.1105254 |
| 361 | Rv0871 | cspB | -3.1091986 |
| 362 | Rv2842c | Rv2842c | -3.1089182 |
| 363 | Rv1611 | trpC | -3.1035643 |
| 364 | Rv0606 | Rv0606 | -3.0992465 |
| 365 | Rv1643 | rplT | -3.0853136 |
| 366 | Rv3049c | Rv3049c | -3.0841708 |
| 367 | Rv1368 | lprF | -3.0821364 |
| 368 | Rv0466 | Rv0466 | -3.0793142 |
| 369 | Rv3046c | Rv3046c | -3.0610368 |
| 370 | Rv0636 | hadB | -3.0609033 |
| 371 | Rv0983 | pepD | -3.059098 |
| 372 | MTB000076 | MTS1082 | -3.0537696 |
| 373 | Rv2248 | Rv2248 | -3.053524 |
| 374 | Rv0642c | mmaA4 | -3.048999 |
| 375 | Rv0571c | Rv0571c | -3.0485687 |
| 376 | Rv3683 | Rv3683 | -3.045284 |
| 377 | Rv1626 | Rv1626 | -3.0407372 |
| 378 | Rv2867c | Rv2867c | -3.0321555 |
| 379 | Rv2477c | Rv2477c | -3.0280125 |
| 380 | Rv2941 | fadD28 | -3.0089047 |
| 381 | Rv1163 | narJ | -2.9730058 |
| 382 | Rv0704 | rplB | -2.9690804 |
| 383 | Rv3914 | trxC | -2.9643416 |
| 384 | Rv1465 | Rv1465 | -2.9589314 |
| 385 | Rv0290 | eccD3 | -2.9490206 |
| 386 | Rv3139 | fadE24 | -2.938962 |
| 387 | Rv3455c | truA | -2.919085 |
| 388 | Rv1645c | Rv1645c | -2.8974183 |
| 389 | Rv0782 | ptrBb | -2.8959744 |
| 390 | Rv3220c | Rv3220c | -2.893275 |
| 391 | Rv0126 | treS | -2.880587 |
| 392 | Rv0153c | ptbB | -2.8739204 |
| 393 | Rv0692 | Rv0692 | -2.87058 |
| 394 | Rv0988 | Rv0988 | -2.8469849 |
| 395 | Rv2198c | mmpS3 | -2.846585 |
| 396 | Rv3708c | asd | -2.8325012 |
| 397 | Rv3249c | Rv3249c | -2.7991006 |
| 398 | Rv0684 | fusA1 | -2.7937715 |
| 399 | Rv1657 | argR | -2.7858322 |
| 400 | Rv0693 | pqqE | -2.7810042 |
| 401 | Rv0559c | Rv0559c | -2.7729316 |
| 402 | Rv3877 | eccD1 | -2.7602134 |
| 403 | Rv0002 | dnaN | -2.7568693 |
| 404 | Rv3443c | rplM | -2.737155 |
| 405 | Rv0479c | Rv0479c | -2.7328978 |
| 406 | Rv0452 | Rv0452 | -2.7324197 |
| 407 | Rv2375 | Rv2375 | -2.7317886 |
| 408 | Rv1248c | Rv1248c | -2.7237267 |
| 409 | Rv0877 | Rv0877 | -2.6837583 |
| 410 | Rv2785c | rpsO | -2.6824555 |
| 411 | Rv1836c | Rv1836c | -2.6820617 |
| 412 | Rv2195 | qcrA | -2.6810896 |
| 413 | Rv2239c | Rv2239c | -2.6774466 |
| 414 | Rv1862 | adhA | -2.675338 |
| 415 | Rv1440 | secG | -2.6742082 |
| 416 | Rv2837c | Rv2837c | -2.6694562 |
| 417 | Rv1801 | PPE29 | -2.6687155 |
| 418 | MTB000061 | mpr5 | -2.6673512 |
| 419 | Rv0315 | Rv0315 | -2.663602 |
| 420 | Rv2909c | rpsP | -2.6629739 |
| 421 | Rv0181c | Rv0181c | -2.6595497 |
| 422 | Rv0808 | purF | -2.6559563 |
| 423 | Rv3747 | Rv3747 | -2.6522098 |
| 424 | Rv0623 | vapB30 | -2.6310413 |
| 425 | Rv0954 | Rv0954 | -2.6297133 |
| 426 | Rv1826 | gcvH | -2.6119883 |
| 427 | Rv2178c | aroG | -2.5995712 |
| 428 | Rv0169 | mce1A | -2.5960054 |
| 429 | Rv1363c | Rv1363c | -2.5872116 |
| 430 | Rv0262c | aac | -2.5501018 |
| 431 | Rv2196 | qcrB | -2.5452309 |
| 432 | Rv1206 | fadD6 | -2.5376463 |
| 433 | Rv0469 | umaA | -2.5255892 |
| 434 | Rv0351 | grpE | -2.499127 |
| 435 | Rv2605c | tesB2 | -2.493807 |
| 436 | Rv3324A | Rv3324A | -2.4937658 |
| 437 | Rv1472 | echA12 | -2.4778452 |
| 438 | Rv1861 | Rv1861 | -2.4609036 |
| 439 | Rv2704 | Rv2704 | -2.4600835 |
| 440 | Rv1095 | phoH2 | -2.4361355 |
| 441 | Rv2387 | Rv2387 | -2.4260721 |
| 442 | Rv3286c | sigF | -2.424395 |
| 443 | Rv3703c | Rv3703c | -2.4216428 |
| 444 | Rv2690c | Rv2690c | -2.4188523 |
| 445 | Rv2587c | secD | -2.4157581 |
| 446 | Rv0440 | groEL2 | -2.415497 |
| 447 | Rv3250c | rubB | -2.4121277 |
| 448 | Rv2631 | Rv2631 | -2.4099727 |
| 449 | Rv0570 | nrdZ | -2.3892858 |
| 450 | Rv0717 | rpsN1 | -2.3884125 |
| 451 | Rv0694 | lldD1 | -2.3785667 |
| 452 | Rv0903c | prrA | -2.3782897 |
| 453 | Rv0805 | Rv0805 | -2.36885 |
| 454 | Rv3223c | sigH | -2.350495 |
| 455 | Rv2995c | leuB | -2.3463635 |
| 456 | Rv3814c | Rv3814c | -2.3456204 |
| 457 | Rv1906c | Rv1906c | -2.3438044 |
| 458 | Rv2535c | pepQ | -2.3422444 |
| 459 | Rv0170 | mce1B | -2.3365357 |
| 460 | Rv1796 | mycP5 | -2.3356552 |
| 461 | Rv1223 | htrA | -2.332401 |
| 462 | Rv3416 | whiB3 | -2.329815 |
| 463 | Rv1435c | Rv1435c | -2.3177435 |
| 464 | Rv3842c | glpQ1 | -2.3075674 |
| 465 | Rv2858c | aldC | -2.296333 |
| 466 | Rv0107c | ctpI | -2.2917602 |
| 467 | Rv3145 | nuoA | -2.289738 |
| 468 | Rv3778c | Rv3778c | -2.285846 |
| 469 | Rv3704c | gshA | -2.2819886 |
| 470 | Rv2129c | Rv2129c | -2.270589 |
| 471 | Rv0476 | Rv0476 | -2.245408 |
| 472 | Rv3291c | lrpA | -2.2329617 |
| 473 | Rv1789 | PPE26 | -2.2231455 |
| 474 | Rv0057 | Rv0057 | -2.217819 |
| 475 | Rv2299c | htpG | -2.2072616 |
| 476 | Rv2703 | sigA | -2.204511 |
| 477 | Rv2102 | Rv2102 | -2.191283 |
| 478 | Rv3681c | whiB4 | -2.1899545 |
| 479 | Rv2500c | fadE19 | -2.1850843 |
| 480 | Rv1162 | narH | -2.1769197 |
| 481 | Rv3280 | accD5 | -2.1565285 |
| 482 | Rv1391 | dfp | -2.1557584 |
| 483 | Rv2501c | accA1 | -2.1364665 |
| 484 | Rv2223c | Rv2223c | -2.1293 |
| 485 | Rv1017c | prsA | -2.1228745 |
| 486 | Rv2444c | rne | -2.1113365 |
| 487 | Rv2878c | mpt53 | -2.0886576 |
| 488 | Rv1306 | atpF | -2.0810251 |
| 489 | Rv0451c | mmpS4 | -2.0457208 |
| 490 | Rv0934 | pstS1 | -2.0456784 |
| 491 | Rv1310 | atpD | -2.0329978 |
